# Preload insufficiency as common denominator of exertional dyspnoea in distinct post-COVID phenotypes

**DOI:** 10.1101/2025.05.26.25328135

**Authors:** G Oruqaj, K Lo, K Krüger, P Bauer, L Laufer, K Milger, C Tabeling, I Pink, Z Rako, N Kremer, S Yildiz, J Behr, M Hecker, M Kaya, S Kuhnert, HH Krämer-Best, U Matt, S Herold, S Oberwinkler, J Wilhelm, M Witzenrath, T Welte, N Weissmann, HA Ghofrani, F Grimminger, W Seeger, K Tello, N Sommer

## Abstract

**Introduction:** The underlying causes of exertional dyspnoea and exercise limitation in post-COVID syndrome remain uncertain. We performed deep-phenotyping of post-COVID patients to evaluate limitations of ventilation, gas exchange and cardiopulmonary circulation in a multicentre, cross-sectional study.

**Methods:** The dyspnoea index and aerobic exercise performance (peakVO_2_) were determined by questionnaires and cardiopulmonary exercise testing, respectively, in a cohort of 86 post-COVID patients and 12 controls. Lung function, gas exchange and ventilation-perfusion mismatch were evaluated. Cardiac parameters were measured by echocardiography and, in a subgroup, systemic vascular characteristics by pulse wave analysis.

**Results:** Post-COVID patients showed low ventilation at peak exercise [VE(peak)], ventilatory inefficiency, low right heart dimensions and low basal oxygen uptake. In a multivariate regression analysis, ventilatory parameters – high breathing frequency at peak exercise (β=0.15, p=0.004) and low forced expiratory volume in 1 s (β=–0.33, p=0.007) – and right atrial end-systolic area index (RA ESAi; β=–0.34, p<0.001) were independent predictors of dyspnoea, while low VE(peak) (β=0.46, p<0.001) and low aerobic capacity (β=0.51, p<0.001) independently predicted low peakVO_2_. Low RA ESAi was associated with a low diffusion coefficient (r=0.36), low end-tidal pCO_2_ (r=0.39) and high heart rate (r=–0.31). Subgroup analysis of patients showed specific associations between dyspnoea and diastolic and bronchial function, low blood pressure, hyperventilation or oxygen uptake.

**Conclusion:** Preload insufficiency associated with gas exchange disturbances contributes to the sensation of dyspnoea in post-COVID patients, as well as ventilatory limitations, while peakVO_2_ was predominantly associated with aerobic capacity. Three phenotypes were defined, indicating the need for tailored interventions.

**Graphical Abstract:** 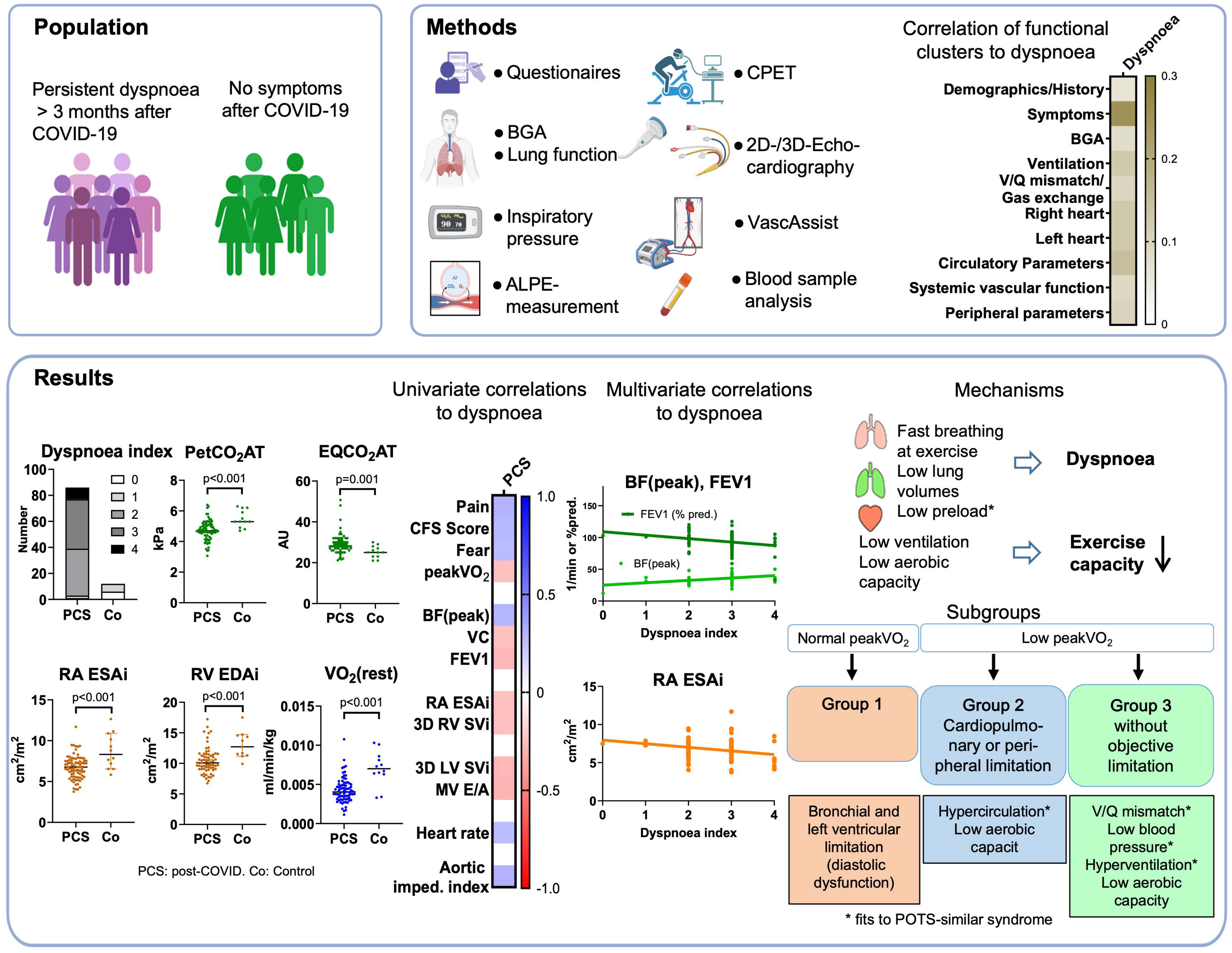

## Introduction

The global COVID-19 pandemic has resulted in long-term impacts of SARS-CoV-2 infection, extending far beyond the acute phase. Among these impacts, post-acute sequelae of COVID-19, referred to as “post-COVID” if symptoms persist beyond 3 months after acute infection, affect millions of patients worldwide [1–3]. Dyspnoea and exercise limitation, a cardinal symptom of post-COVID, persists in a substantial proportion of patients, irrespective of the severity of the initial infection [4, 5]. Other symptoms frequently include cardiac symptoms, such as tachycardia, and neurological symptoms, such as autonomic dysfunction with postural tachycardia (POTS) and post-exertional malaise similar to myalgic encephalomyelitis/chronic fatigue syndrome (ME/CFS) [6–8]. Importantly, symptoms vary among patients and different symptom clusters have been identified in clinical studies [9].

Despite its high prevalence, the mechanisms underlying exertional dyspnoea in post-COVID patients remain poorly understood and are likely multifactorial involving ventilatory, cardiac, vascular and psychological factors [10, 11]. Invasive cardiopulmonary exercise testing (CPET) in post-COVID patients has revealed ventilatory inefficiency (i.e. hyperventilation) and decreased peripheral oxygen extraction at preserved cardiac output as a cause of reduced peak oxygen consumption (peakVO_2_) in post-COVID patients [10], while other studies suggest that deconditioning is the main factor in low peakVO_2_ [12]. Moreover, pulmonary perfusion disturbances, ventilation-perfusion (V/Q) mismatch, and increased alveolar dead space have been observed even months after infection, although the relevance for dyspnoea and exercise limitation has been questioned [4, 10, 13, 14]. Pulmonary hypertension (PH) and right ventricular (RV) dysfunction, often observed in hospitalised COVID-19 patients, have been linked to worse outcomes and may persist in post-COVID recovery, contributing to reduced exercise capacity [15]. However, Singh et al. [10]. reported low biventricular filling pressures in post-COVID patients with exercise limitation, and POTS, which is a frequent condition in a specific subpopulation of post-COVID patients, is associated with a chronic low cardiac preload state (“RV preload insufficiency”) leading to dyspnoea and exercise intolerance [13]. Dyspnoea was also linked to ongoing myocardial inflammation and left heart dysfunction [16], as well as systemic endothelial dysfunction [17].

We aimed to investigate cardiopulmonary factors associated with post-COVID-related dyspnoea and exercise limitation. This study integrates assessments of CPET, lung function tests, gas exchange, V/Q mismatch and echocardiographic measurements and, in a subgroup, right heart catheterisation and systemic vascular characterisation to investigate cardiopulmonary limitations in post-COVID patients. By correlating these findings with patient-reported symptoms and exercise performance, we aim to provide a comprehensive understanding of the mechanisms driving post-COVID exertional limitation in specific patient sub-populations and identify potential therapeutic targets.

## Methods

### Study design and study population

This multicentre, cross-sectional study titled “Pulmonary Vascular Dysfunction as a Cause of Persistent Exertional Dyspnoea Following COVID-19” (PulmVasC) was conducted between 02/2022 and 02/2024 across four German tertiary care centres, enrolling 86 patients with post-COVID syndrome and 12 healthy controls (NCT05374577). The study was approved by the local ethics committee (Justus-Liebig University Giessen, reference number AZ 28/22). All patients had a confirmed SARS-CoV-2 infection and unexplained dyspnoea persisting or developing after more than 3 months after the acute infection according to the World Health Organization definition of the post-COVID condition. Controls were individuals with a documented SARS-CoV-2 infection and no chronic symptoms. A standardised outpatient protocol was performed including symptom questionnaires, imaging, functional tests and, in a subgroup of patients, right heart catheterisation and biomarker sampling (see SI Methods). Symptom questionnaires, including the Chandler Fatigue Scale, were used to assess fatigue, mental health, and functional limitations. Fasting venous blood sampling was performed for routine lab testing and further biomarker analyses.

### Parameter measurements

**Pulmonary function** testing included body plethysmography and diffusion capacity (DLCO) measurement following international guidelines, with results expressed as % predicted.

**Ventilation-perfusion (V/Q) mismatch** was evaluated using the Automatic Lung Parameter Estimator (ALPE2, Mermaid Care A/S, Stenlose, Denmark), which estimates ΔpO_₂_ and ΔpCO_₂_ from exhaled gases (pCO_2_ and pO_2_) and capillary blood gases during controlled FiO_₂_ steps [18].

**CPET** was conducted using semi-recumbent ergometry with ramp protocols individualised by physical activity level. Gas exchange, ventilatory efficiency and peakVO_₂_ were determined breath-by-breath in accordance with current international guidelines [19]. Exercise echocardiography was performed in parallel to assess left and right heart function. **Comprehensive echocardiography** (2D and 3D) was performed per current standards, with volumetric and strain analyses indexed to body surface area [20].

**Pulse pressure waveform analysis** was used to assess central and peripheral vascular function, including pulse wave velocity, the augmentation index, and ejection time via oscillometry (VascAssist2, iSYMED GmbH, Butzbach, Germany) [21].

**Right heart catheterisation** was conducted in a clinically indicated subset, measuring mean pulmonary arterial pressure (mPAP), PAWP, CO, and PVR using Fick and thermodilution methods, as described previously [20].

### Statistical analysis

Statistical analysis included t-tests, linear regression, and Fisher’s r-to-z transformation for grouped correlation analysis. Full methods are detailed in the Supplementary Information.

## Results

### Basic characteristics of the PulmVasC patient cohort

A total of 98 participants were included in the analysis, comprising 86 patients and 12 controls with a median age of 44 and 37 years, respectively (Table 1). At the time of investigation, patients reported a median of 7 out of a maximum of 21 questionnaire-related symptoms, whereas controls reported none. Patients, but not controls, experienced an increase in dyspnoea following SARS-CoV-2 infection, with a median time since infection of 323 and 448 days, respectively. Post-COVID symptom scores highlighted a higher symptom burden among patients compared to controls (Table 1 and Supplementary Table S1). PeakVO_₂_ in the overall patient group was reduced to 86% of the predicted value. Vital parameters and blood gas analyses revealed no pathological values or significant differences between patients and controls (Table 1).

**Table 1:**
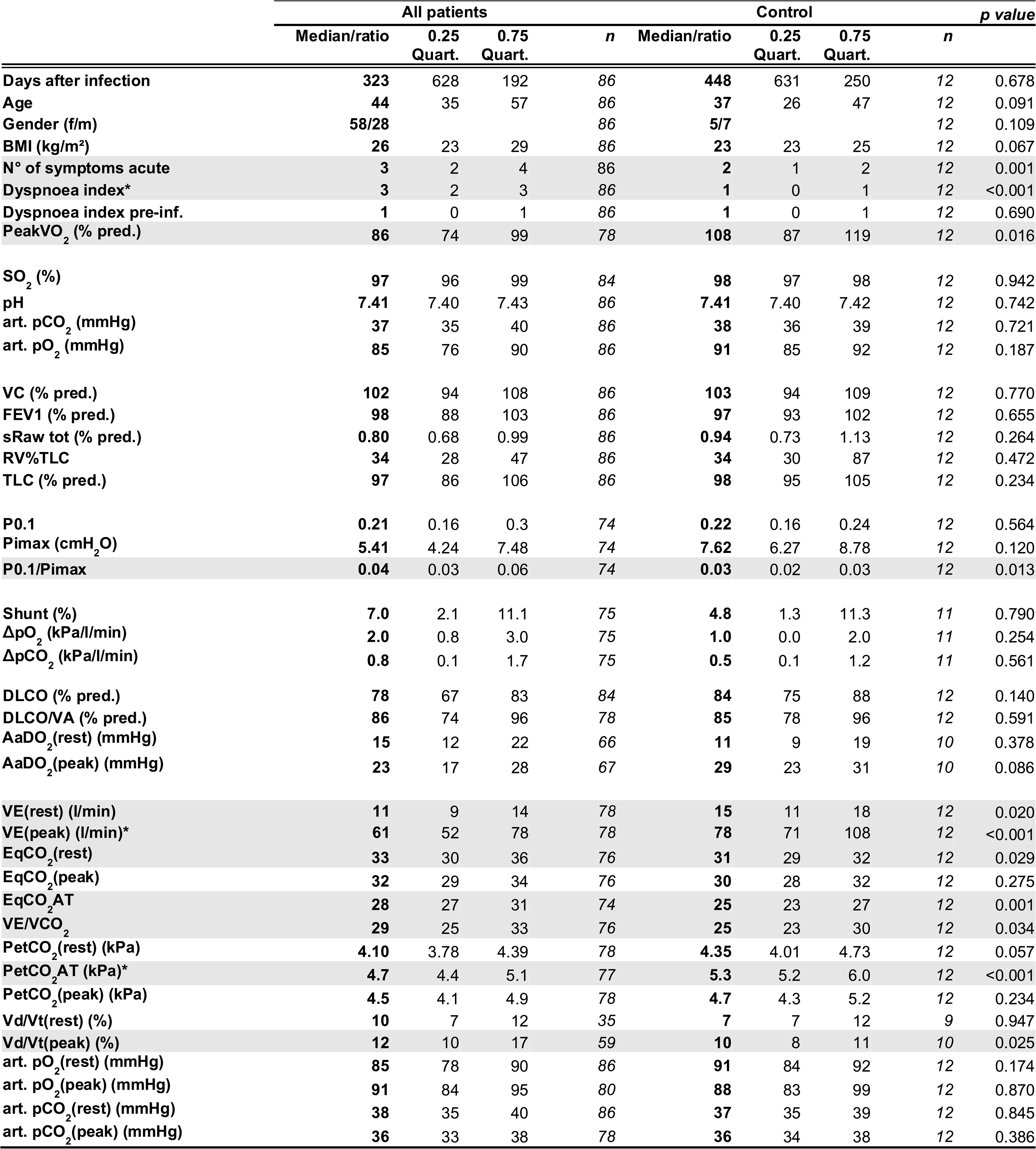
Demographic characteristics. symptom load. blood gases. ventilatory parameters. V/Q mismatch and gas exchange parameters of all patients and control subjects provided as median and interquartile range. Unpaired Student‘s t-test or Mann-Whitney U test were performed as appropiate. Parameters with p<0.05 are highlighted in gray. * significant differences (adjusted to false discovery rate) *f/m: female/male, BMI: body mass index, N°: number. pre-inf.: pre-infection, peakVO2: oxygen uptake at peak exercise, pred.: predicted, SO2: oxygen saturation, art. pCO2: partial pressure of carbon dioxide, art. pO2: partial pressure of oxygen, VC: vital capacity, FEV1: forced expiratory volume in 1 s, sRaw tot: total specific resistance, RV%TLC: residual volume in % total lung capacity, TLC: total lung capacity, P0.1: airway occlusion pressure, Pimax: maximal inspiratory pressure, P0.1/Pimax: airway occlusion pressure/maximal inspiratory pressure,* Δ*pO2: measure for low V/Q area,* Δ*pCO2: measure for high V/Q areas, DLCO: diffusing capacity for carbon monoxide, DLCO/VA: diffusing capacity/alveolar volume, AaDO2: alveolar-arterial oxygen gradient, VE: ventilation, EqCO2: ventilatory equivalent of carbon dioxide, EqO2: ventilatory equivalent of oxygen, VE/VCO2: ventilation/carbon dioxide production, PetCO2: end-tidal carbon dioxide tension, VDc/VT: ratio of physiologic dead space over tidal volume*

### Post-COVID patients showed ventilatory inefficiency, reduced right heart filling and low aerobic metabolism

We measured approximately 150 parameters by different non-invasive cardiopulmonary and vascular measurement methods characterising different functional groups of the cardiopulmonary system (Fig. 1a). Patients with post-COVID syndrome showed higher dyspnoea and lower performance and ventilation at maximal exercise [VE(peak)] in CPET (Fig. 1b i–iii, Table 1, Supplementary Table S1). Moreover, the partial pressure of end-tidal carbon dioxide at the anaerobic threshold (PetCO_2_AT) was decreased (Fig. 1b iv) and the ventilator equivalent for carbon dioxide at the anaerobic threshold (EQCO_2_AT) was increased (Fig. 1b v, Table 1) in post-COVID patients compared with controls (Co). These results indicate an augmented ventilatory drive and/or V/Q mismatch, which is in accordance with high dead-space ventilation (see Vd/Vt, Table 1) or areas with high V/Q (see Table 1). Interestingly, the right atrial end-systolic area index (RA ESAi) and right ventricular end-diastolic area index (RV EDAi) was reduced in post-COVID patients vs. controls (Fig. 1b vi–vii, Table 2), supporting a component of low Q contributing to V/Q mismatch. Moreover, oxygen uptake at rest, VO_2_(rest), was lower in post-COVID patients compared to controls (Fig. 1b viii). Parameters were grouped according to functional clusters and their correlation coefficient was averaged for each cluster to understand the relevance of different functional clusters for the development of dyspnoea or low peakVO_2_ (Fig. 1c i). Dyspnoea was associated across all patients with high symptom load, and weakly with ventilatory, cardiac and circulatory parameters (Fig. 1c ii). PeakVO_2_ showed associations with left heart, circulatory, systemic vascular and peripheral parameters (Fig. 1c ii).

**Fig. 1.**
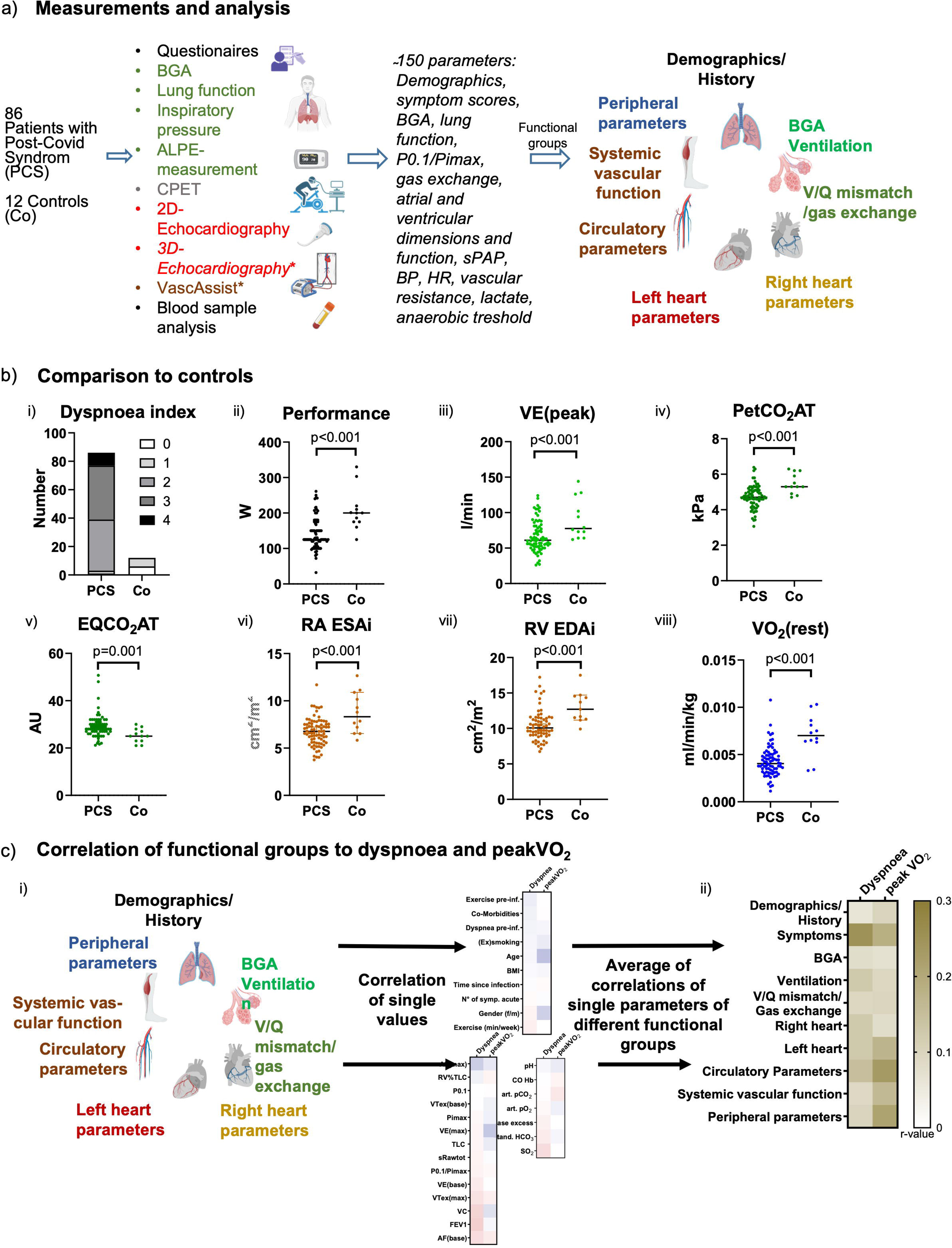
Overview of the PulmVasC multicentre study design and findings in patients with post-COVID syndrome. **a)** Schematic overview of the PulmVasC study. A total of 86 patients with post-COVID syndrome and 12 healthy controls underwent a standardised sequence of clinical assessments. All patients presented with persistent dyspnoea more than 3 months following SARS-CoV-2 infection. Across all assessments, approximately 150 parameters were collected that can be categorised into different functional groups. **b)** Comparison between post-COVID patients and controls. Significant differences were found for the dyspnoea index (i), performance (ii), ventilation at peak exercise [VLJE(peak)] (iii), end-tidal CO_₂_ partial pressure at the anaerobic threshold (PetCO_₂_AT) (iv), ventilatory equivalent for CO_₂_ at the anaerobic threshold (EqCO_₂_AT) (v), right atrial end-systolic area index (RA ESAi) (vi), right atrial end-diastolic area index (RA EDAi) (vii), and resting oxygen uptake [VO_2_(rest)] (viii). Parameters of post-COVID patients and control participants were compared by unpaired Student’s t-test or the U Mann–Whitney test, respectively, and significances were adjusted for the false discovery rate. **c)** Correlation analysis between functional groups and dyspnoea or peakVO_2_. Individual correlations between each parameter and dyspnoea or peakVO_₂_ were computed. Fisher’s r-to-z transformation was applied to each correlation coefficient, and the transformed absolute z-values of parameters belonging to a specific functional group were averaged and re-transformed to r-values, which are displayed in brown. Grouping of parameters to a specific functional group is provided in the Supplementary Figures. P0.1/Pimax, respiratory drive index; BF, breathing frequency; EqCO_₂_, ventilatory equivalent for CO_₂_; sPAP, systolic pulmonary artery pressure; BP, blood pressure; HR, heart rate; BGA, blood gas analysis; VLJE(peak), ventilation at peak exercise; PetCO_₂_AT, end-tidal CO_₂_ partial pressure at anaerobic threshold; RA ESAi, right atrial end-systolic area index; RA EDAi, right atrial end-diastolic area index; VO_₂_(rest), oxygen uptake at rest.

**Table 2:**
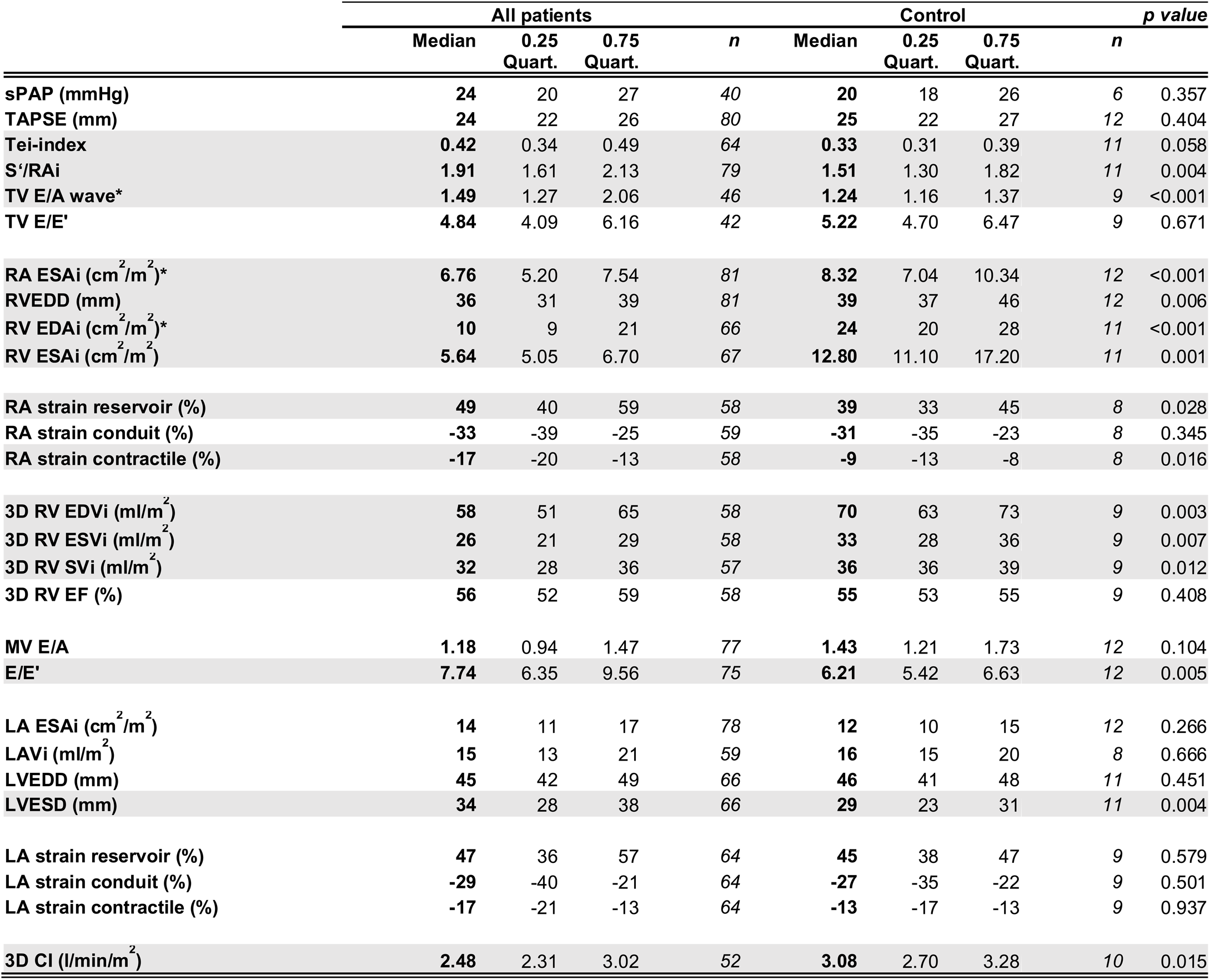
Parameters of right and left heart of all patients and control subjects provided as median and interquartile range. Unpaired Student‘s t-test or Mann-Whitney U test were performed as appropiate. Parameters with p<0.05 are highlighted in gray. * significant differences (adjusted to false discovery rate). *sPAP: systolic pulmonary artery pressure, TAPSE: tricuspid annular plane systolic excursion, Tei-index: myocardial performance index, S‘/RAi: systolic contraction velocity/right atrial index, TV: tricuspid valve, E/A wave: early to late diastolic transtricuspid flow velocity, E/E‘: ratio of the E wave velocity to myocardial movement velocity, RA: right atrium, ESAi: end-systolic area index, RV: right ventricular, EDD: end-diastolic diameter, EDAi: end-diastolic area index, ESAi: end-systolic area index, EDVi: end-diastolic volume index, ESVi: end-systolic volume index, SVi: stroke volume index, EF: ejection fraction, LA: left atrial, LAVI: left atrial volume index. CI: Cardiac index, LVEDD: left ventricular end-diastolic diameter, LVESD: left ventricular end-systolic diameter*

### Association of ventilatory but not gas exchange parameters with dyspnoea

We then investigated the correlations of the single parameters of the functional clusters with dyspnoea or peakVO_2_ in post-COVID patients. Parameters with an r-value > 0.25 or < –0.25 were used for further analysis and are shown compared to the controls (Fig. 2). As expected, dyspnoea was positively associated with symptoms and negatively associated with peakVO_2_ (Fig. 2a), while peakVO_2_ was associated with age, female gender, performance and low dyspnoea (Fig. 2b). Importantly, a high respiratory rate at peak exercise [BF(peak)], low vital capacity (VC in % predicted) and low forced expiratory volume in 1 s (FEV1 in % predicted) were associated with high levels of dyspnoea in post-COVID patients but not controls (Fig. 2 a, c i). In contrast, markers of V/Q mismatch (Shunt, ΔpO_2_, and ΔpCO_2_) and gas exchange (pO_2_, DLCO, and AaDO_2_) were not correlated with dyspnoea (Supplementary Fig. 1d), although shunting, areas of low V/Q (represented by ΔpO_₂_), and areas of high V/Q (represented by ΔpCO_₂_) were relatively high in post-COVID patients compared to controls and published normal values (22) (Table 1). Accordingly, the diffusion capacity for carbon monoxide (DLCO, % predicted) was below the normal range (76–140%) in approximately 50% of the patients, and levels of partial pressure for oxygen (pO_₂_) were relatively low at rest (Table 1). As arterial pO_2_ increased adequately during exercise and no abnormalities were observed in the alveolar-arterial oxygen difference during exercise [AaDO_₂_(peak)] (Table 1), gas exchange impairments are probably compensated during exercise and, in general, do not contribute to the sensation of dyspnoea. Thus, shallow breathing but not hypoxaemia or diffusion limitation may contribute to the sensation of dyspnoea during exercise in post-COVID patients and result in signs of V/Q mismatch. In contrast, high peakVO_2_ was associated with high maximal ventilation [VE(peak)] and alveolar-arterial difference in pO_2_ (AaDO_2_), which may be affected by the level of effort (Fig. 2b, d i).

**Fig. 2.**
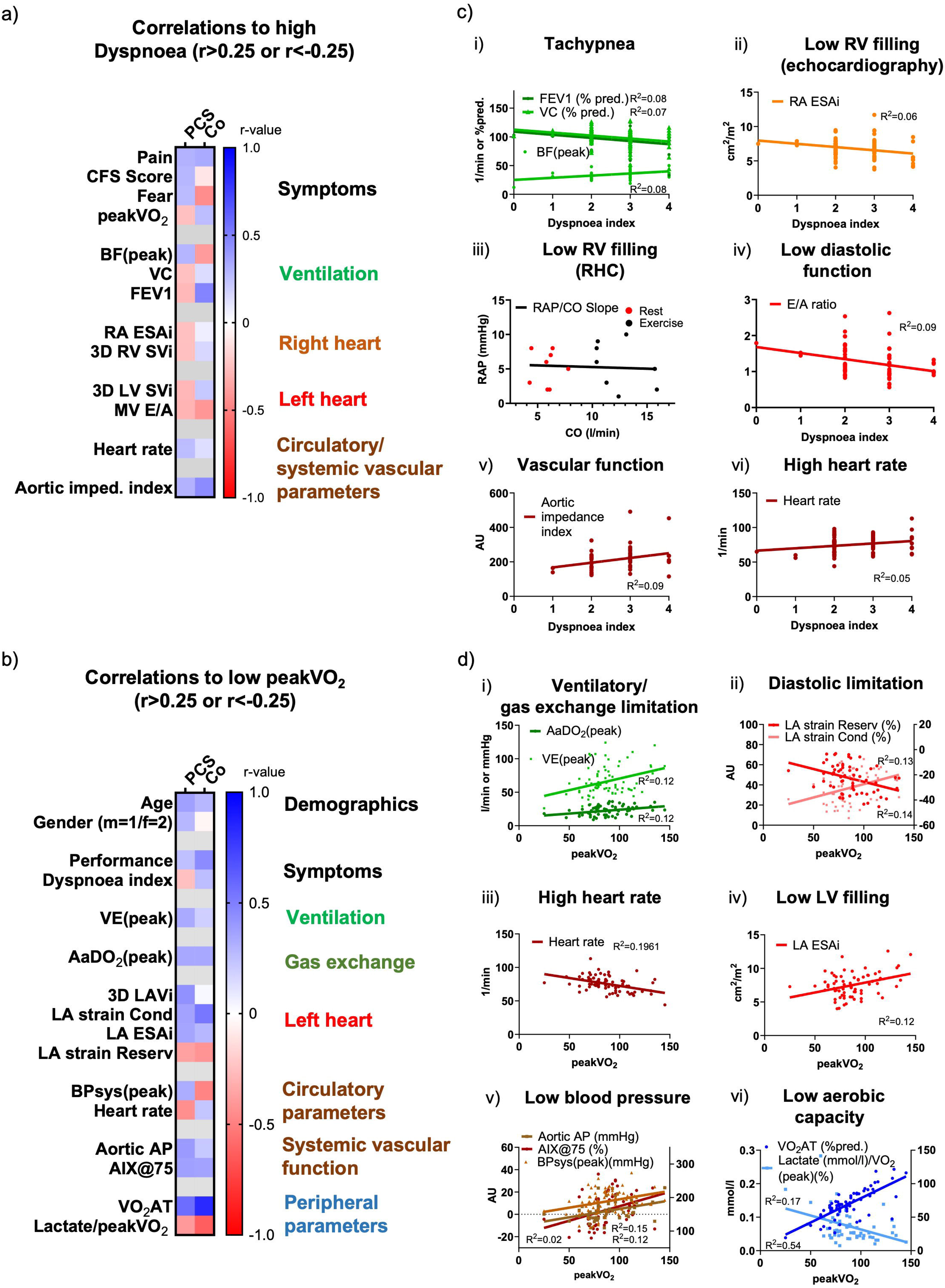
Correlations of dyspnoea and aerobic exercise capacity (peakVO_₂_) with demographic, symptomatic, cardiopulmonary, and vascular parameters in the PulmVasC cohort. **a)** Correlations between dyspnoea or **b)** peakVO_2_ with measurement parameters demonstrating r-values > 0.25 or < –0.25 in a simple linear regression analysis. **c)** Selected linear regression analyses for dyspnoea: i) breathing frequency at peak exercise (BF(peak), vital capacity (VC), and forced expiratory volume in 1 s (FEV_₁_); ii) right atrial end-systolic area index (RA ESAi); iii) relationship of cardiac output (CO) to right atrial pressure (RAP) determined by right heart catheterisation; iv) mitral valve E/A ratio (MV E/A); v) aortic impedance index; vi) resting heart rate. **d)** Selected linear regression analyses for peak VO_₂_: i) minute ventilation at peak exercise [VLJE(peak)] and alveolar-arterial oxygen difference at peak exercise [(AaDO_₂_(peak)]; ii) left atrial (LA) strain; iii) LA end-systolic area index (LA ESAi); iv) resting heart rate; v) aortic augmentation pressure (aortic AP), augmentation index at 75 bpm (AIX75%), and systolic blood pressure at peak exercise [(BPsys(peak)]; vi) oxygen uptake at the anaerobic threshold (VO_₂_AT). CFS, Chronic fatigue score; peakVO_₂_, peak oxygen uptake; BF(peak), breathing frequency at peak exercise; VC, vital capacity; FEV_₁_, forced expiratory volume in 1 s; RA ESAi, right atrial end-systolic area index; 3D RV SVi, right ventricular stroke volume index (3D echocardiography); MV E/A, mitral valve early/late diastolic filling velocity ratio; 3D LV SVi, left ventricular stroke volume index (3D echocardiography); VLJE peak, minute ventilation at peak exercise; AaDO_₂_(peak), alveolar-arterial oxygen difference at peak exercise; LA strain, left atrial strain (reservoir and conduit phases); LA ESAi, left atrial end-systolic area index; aortic AP, aortic augmentation pressure; AIX75%, augmentation index adjusted to a heart rate of 75 bpm.

### Right heart parameters indicate preload insufficiency in post-COVID

Right ventricular (RV) and right atrial (RA) parameters revealed no evidence of pulmonary hypertension (PH) or RV overload, either at rest (Table 2, Supplementary Table S3) or during exercise (Supplementary Table S4). However, the median Tei index was elevated (normal value: 0.28 ± 0.04) (23). RV volumes were reduced compared to established normal ranges reported in the literature (end-diastolic volume [EDVi]: 75 ± 12 mL/m^2^; end-systolic volume [ESVi]: 33 ± 7 mL/m^2^) (24), and RA ESAi, as well as RV EDAi, were significantly decreased compared to controls (Fig. 1b vi, vii, Table 2). Additionally, RA strain during the reservoir phase was elevated and in the high range compared to published normal values (average value: 44%; 95% CI: 25–63%) (25) suggesting low right atrial pressure. Importantly, the severity of dyspnoea was associated with reduced right heart volumes, in particular, low RA ESAi, and function, both at rest (Fig. 2c ii, Supplementary Fig. 1e) and during exercise (Supplementary Table S4).

We hypothesised that the association of reduced heart volumes may indicate right heart preload insufficiency. We analysed right heart catheter (RHC) data from eight post-COVID patients in whom RHC was performed for clinical reasons. In these patients, no evidence of PH was observed, either at rest or during exercise (Supplementary Table S5). However, the limited rise in RA pressure during exercise may suggest preload insufficiency, as previously defined (26, 27). Indeed, we observed a negative RAP/CO-slope (Fig. 2c iii), in contrast to the positive RAP/CO-slopes found in healthy individuals [28, 29].

### Exercise capacity is associated with diastolic dysfunction and systemic circulatory parameters

Although left ventricular parameters did not show pathological values (Table 2, Supplementary Table S6), both high dyspnoea and low peakVO_2_ were linked to low diastolic function, as characterised by MV E/A or LA strain values in post-COVID patients and controls (Fig. 2a, b, c iv, d ii), indicating mechanisms that were not specifically related to post-COVID syndrome. The association between high peakVO_2_and high LA ESAi (Fig. 2d iii) may indicate efficient LA filling. Importantly, both high dyspnoea and low peakVO_2_ positively correlated with basal heart rate specifically in post-COVID patients but correlated negatively in controls (Fig. 2a, b, c vi, d iv). The correlation between peakVO_2_ and maximal systolic blood pressure [BPsys(peak)] may be a reflection of maximal effort rather than limitation during exercise (Fig. 2b, d v). In a subgroup of 56 patients, pulse wave analysis was performed to determine the central and peripheral systemic vascular parameters (Table 3). Consistent with the observed association between dyspnoea and diastolic dysfunction, a high aortic impedance index, an indicator of aortic stiffness, was associated with greater dyspnoea in post-COVID patients and controls (Fig. 2a, c v).

**Table 3:** Pulse wave analysis and peripheral parameters of all patients and control subjects provided as median and interquartile range. Unpaired Student‘s t-test or Mann-Whitney U test were performed as appropiate. Parameters with p<0.05 are highlighted in gray. * significant differences (adjusted to false discovery rate). *PWV: pulse wave velocity, BP: blood pressure, ABI: Ancle-brachial-index, RER: respiratory exchange ratio, VO2: oxygen uptake, AT: anaerobic*

|  | All patients |  |  |  | Control |  |  |  | p value |
| --- | --- | --- | --- | --- | --- | --- | --- | --- | --- |
|  | Median | 0.25 Quart. | 0.75 Quart. | n | Median | 0.25 Quart. | 0.75 Quart. | n |  |
| Aortic PWV (m/s) | 7.6 | 6.8 | 8.3 | 57 | 6.6 | 6.1 | 7.2 | 10 | 0.028 |
| Brachial BP (mmHg) | 126 | 118 | 137 | 57 | 118 | 113 | 126 | 10 | 0.178 |
| Aortic BP (mmHg) | 107 | 99 | 117 | 57 | 108 | 94 | 114 | 10 | 0.463 |
| Brachial ABI left | 1.16 | 1.11 | 1.22 | 45 | 1.17 | 1.14 | 1.23 | 5 | 0.437 |
| Brachial ABI right | 1.17 | 1.11 | 1.23 | 47 | 1.20 | 1.16 | 1.23 | 9 | 0.974 |
| Brachial PWV left | 9.85 | 9.20 | 11.09 | 44 | 8.95 | 8.66 | 10.23 | 0 | 0.027 |
| Brachial PWV right | 10.53 | 9.43 | 11.39 | 46 | 9.70 | 8.56 | 10.16 | 10 | 0.058 |
| pH, peak | 7.37 | 7.32 | 7.41 | 53 | 7.29 | 7.28 | 7.31 | 12 | 0.002 |
| Lactate (mmol/l)/peakVO<sub>2</sub> (%) | 0.06 | 0.04 | 0.09 | 65 | 0.10 | 0.06 | 0.11 | 10 | 0.070 |
| RER(rest) | 0.82 | 0.77 | 0.93 | 76 | 0.84 | 0.71 | 0.91 | 12 | 0.671 |
| RER(peak) | 1.11 | 1.04 | 1.18 | 76 | 1.17 | 1.13 | 1.22 | 12 | 0.059 |
| VO<sub>2</sub>AT (%max) | 84 | 77 | 88 | 66 | 87 | 83 | 94 | 12 | 0.146 |
| VO<sub>2</sub>AT (%Soll) | 67 | 58 | 82 | 77 | 87 | 81 | 103 | 12 | 0.002 |
| VO<sub>2</sub>(rest) (ml/min/kg)* | 0.004 | 0.003 | 0.005 | 76 | 0.007 | 0.006 | 0.008 | 12 | <0.001 |

### Low exercise capacity is associated with low aerobic capacity

High lactate and low VO_2_AT levels were associated with low peakVO_2_, suggesting a limitation in exercise capacity due to impaired aerobic capacity in post-COVID patients and controls (Fig. 2b, d vi), again suggesting that this is a mechanism not related to post-COVID syndrome.

### Ventilatory parameters, low right heart preload and low aerobic capacity are independent predictors of dyspnoea and/or exercise capacity

Using multivariate linear regression analysis incorporating parameters from the univariate analysis along with potential confounders (age and gender), we found that FEV1, BF(peak) and RA ESAi were independent predictors of dyspnoea (Table 4). Low FEV1 was associated with worse vascular function and low RA ESAi with a low diffusion coefficient, low PetCO_2_AT and a high heart rate (Supplementary Fig. 5). Furthermore, low VE(peak) and VO_2_AT independently predicted exercise capacity. In summary, ventilatory parameters and reduced cardiac filling associated with gas exchange disturbances emerge as key predictors of dyspnoea.

**Table 4:** Multivariate linear regression analysis of dyspnoea and peakVO2 including potential cofounders and signficiant correlations from univariate analysis *VC: vital capacity, FEV1: forced expiratory volume in 1 sec, BF(peak): breathing frequency at peak exercise, RA ESAi: right atrial end-systolic area index, MV E/A: ratio of the early (E-wave) to late (A-wave) ventricular filling velocities at the mitral valve, AaDO2: Alveolar-arterial oxygen tension difference at peak exercise, VE: ventilation, BPsys: systolic blood pressure, LA ESAi: left atrial end-systolic area index, LA: left atrial, VO2AT: oxygen uptake at the anerobic treshold, peakVO2: oxygen uptake at peak exercise, aortic, AP: aortic arterial pressure, AIX@75: heart rate-corrected augmentation index*

|  | Variable | Estimate | Standard error | 95% CI (asymptotic) | P value | P value summary |
| --- | --- | --- | --- | --- | --- | --- |
| Dyspnoea | Intercept | 0.00 | 0.01 | -0.02 to 0.01 | 0.83 | ns |
|  | Age | 0.11 | 0.07 | -0.03 to 0.24 | 0.13 | ns |
|  | Gender (f/m) | -0.14 | 0.06 | -0.25 to -0.025 | 0.02 | * |
|  | Heart rate | 0.12 | 0.06 | -0.01 to 0.24 | 0.07 | ns |
|  | VC (% pred.) | 0.08 | 0.12 | -0.15 to 0.31 | 0.51 | ns |
|  | FEV1 (% pred.) | -0.33 | 0.12 | -0.57 to -0.09 | 0.01 | ** |
|  | Bf(peak) | 0.15 | 0.05 | 0.048 to 0.26 | 0.00 | ** |
|  | RA ESAi (cm<sup>2</sup>/m<sup>2</sup>) | -0.34 | 0.05 | -0.44 to -0.24 | <0.0001 | **** |
|  | MV E/A | -0.10 | 0.08 | -0.25 to 0.06 | 0.23 | ns |
|  | Aortic impedance index | 0.04 | 0.06 | -0.07 to 0.15 | 0.49 | ns |
| PeakVO<sub>2</sub> | Intercept | -17.29 | 37.18 | -94.41 to 59.82 | 0.65 | ns |
|  | Age | 0.24 | 0.12 | -0.02 to 0.50 | 0.06 | ns |
|  | Gender (f/m) | 10.86 | 5.93 | -1.44 to 23.16 | 0.08 | ns |
|  | Heart rate | 0.07 | 0.21 | -0.37 to 0.50 | 0.75 | ns |
|  | AaDO<sub>2</sub>(peak) (mmHg) | 0.08 | 0.23 | -0.40 to 0.56 | 0.73 | ns |
|  | VE(peak) (l/min) | 0.46 | 0.09 | 0.27 to 0.65 | <0.0001 | **** |
|  | BPsyst(peak) (mmHg) | 0.05 | 0.06 | -0.07 to 0.16 | 0.40 | ns |
|  | LA ESAi | 0.66 | 1.11 | -1.65 to 2.96 | 0.56 | ns |
|  | LA strain reserv (%) | -0.29 | 0.27 | -0.85 to 0.28 | 0.31 | ns |
|  | LA strain cond (%) | -0.43 | 0.35 | -1.15 to 0.30 | 0.24 | ns |
|  | VO<sub>2</sub>AT (% pred.) | 0.51 | 0.10 | 0.29 to 0.72 | <0.0001 | **** |
|  | Aortic AP (mmHg) | 0.74 | 0.70 | -0.72 to 2.20 | 0.30 | ns |
|  | AIX@75 (%) | -0.37 | 0.40 | -1.20 to 0.47 | 0.37 | ns |

### Subgroup-specific associations of dyspnoea with cardiovascular dysfunction, low blood pressure and hyperventilation

To further investigate whether the causes of dyspnoea and exercise limitation are different between patients with normal peakVO_2_ and low peakVO_2_ (limited by objective factors or effort), we stratified patients into subgroups based on their CPET characteristics (Fig. 3a, Supplementary Table S1): Group 1 – patients reaching the expected peakVO_2_; Group 2 – patients with diminished peakVO_2_ but objective cardiopulmonary limitations (indicating maximal effort); and Group 3 – patients without objective cardiopulmonary limitations (indicating sub-maximal effort). In particular, patient Group 2 differed from patient Groups 1 and 3 with regard to lower PetCO_2_, higher arterial pO_2_(peak), higher HR(rest and peak), and lower VO_2_AT, while Group 3 showed lower blood pressure values compared to the other groups.

**Fig. 3.**
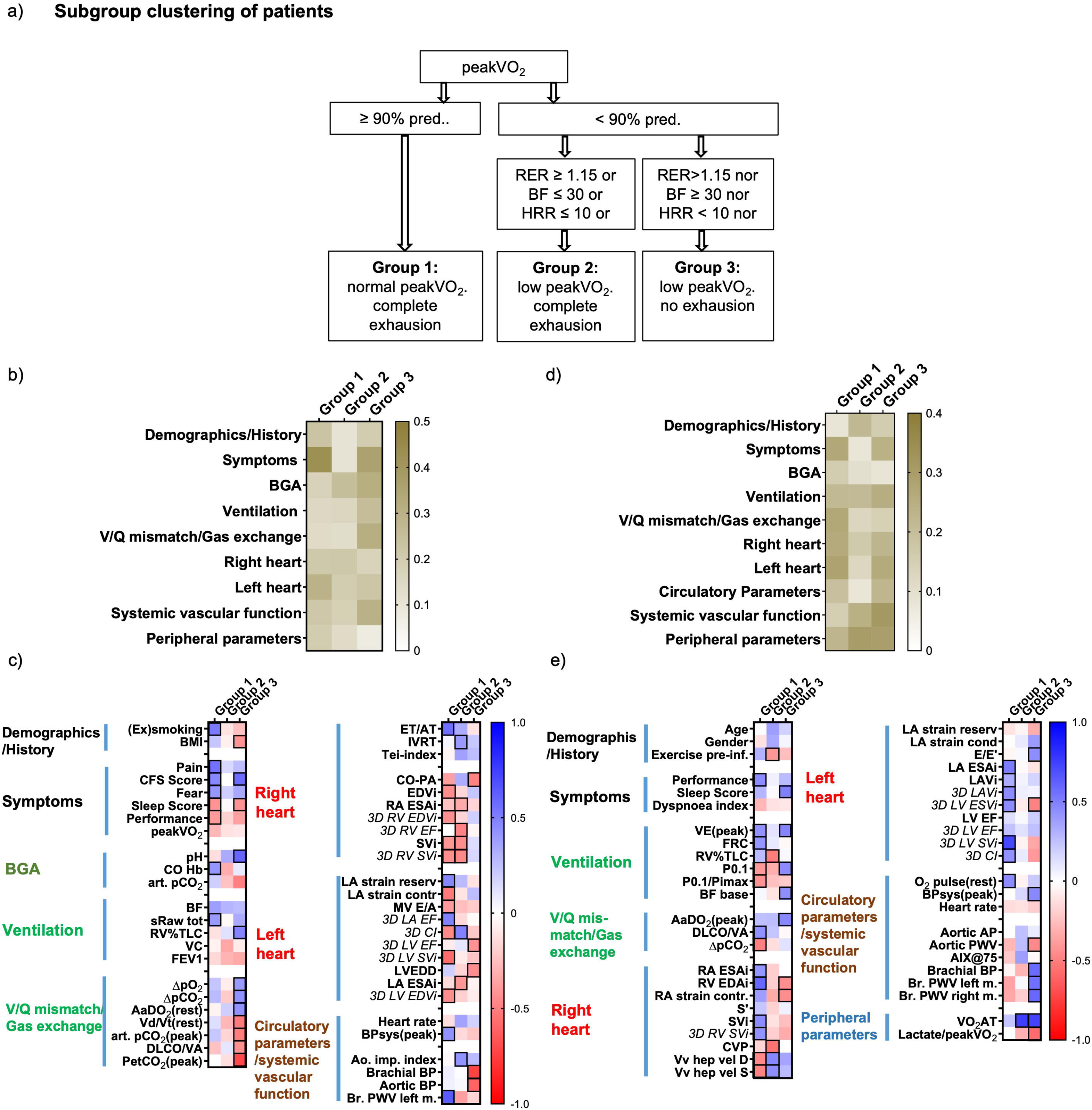
Correlations between dyspnoea and peak VO_2_ with cardiopulmonary parameters across patient subgroups. **a)** Patients were stratified into three subgroups based on their cardiopulmonary exercise testing (CPET) performance: Group 1 reached expected peakVO_₂_ values (normal exercise capacity); Group 2 did not reach expected peakVO_2_ values but showed objective ventilatory, cardiac or metabolic limitations; and Group 3 exhibited none of the above objective limitations. **b)** Correlation between different parameters of functional clusters and dyspnoea within each subgroup. Functional clusters were defined as illustrated in the Supplementary Figures. For each cluster, Fisher’s r-to-z transformation was applied to individual correlation coefficients. The absolute z-values were averaged and re-transformed to r-values, shown in brown. **c)** Subgroup-specific correlations between dyspnoea and individual parameters. Only parameters with correlation coefficients (r) > 0.4 or < –0.4 in a simple linear regression analysis are shown. Correlations of all parameters are shown in Supplementary Fig. 1 and 2. **d)** Correlations between different parameters of functional clusters and peakVO_₂_ within patient subgroups. Functional clusters were defined as described in the Supplementary Figures. For each cluster, Fisher’s r-to-z transformation was applied to individual correlation coefficients. The absolute z-values were averaged within each cluster and re-transformed to r-values, shown in brown. **e)** Subgroup-specific correlations between peakVO_₂_ and individual parameters. Only parameters with correlation coefficients (r) > 0.4 or < –0.4 in a simple linear regression analysis are shown. Correlations of all parameters are shown in Supplementary Fig. 3 and 4. VLJE(peak), minute ventilation at peak exercise; BMI, body mass index; CFS, chronic fatigue score; pred. predicted; peakVO_₂_, peak oxygen consumption; RER, respiratory exchange ratio; CO Hb, carboxyhaemoglobin; art. pCO_₂_, arterial partial pressure of carbon dioxide; ΔpO_₂_, oxygen partial pressure difference due to low V/Q mismatch; ΔpCO_₂_, oxygen partial pressure difference due to high V/Q mismatch; AaDO_₂_(base)/(peak), alveolar-arterial oxygen difference at rest/peak exercise; VLJd/VLJt base, dead space-to-tidal volume ratio at rest; DLCO/VA, diffusing capacity per unit alveolar volume (transfer coefficient); PetCO_₂_(peak), end-tidal CO_₂_ at peak exercise; BF(peak), breathing frequency at peak exercise; RV%TLC, residual volume as percentage of total lung capacity; sRaw tot, total specific airway resistance; VC, vital capacity; FEV_₁_, forced expiratory volume in 1 s; IVRT, isovolumic relaxation time; ET/AT, ejection time to acceleration time ratio; CO-PA, cardiac output measured in the pulmonary artery; EDVi, end-diastolic volume index; 3D RV EDVi, right ventricular end-diastolic volume index (3D echo); 3D RV EF, right ventricular ejection fraction (3D echo); SVi, stroke volume index; RA ESAi, right atrial end-systolic area index; 3D RV SVi, right ventricular stroke volume index (3D echo); LA strain reserve, left atrial reservoir strain; 3D LA EF, left atrial ejection fraction (3D echo); 3D CI, cardiac index (3D echo); LVEDD, left ventricular end-diastolic diameter; 3D LV EF, left ventricular ejection fraction (3D echo); LA strain contr, left atrial contractile strain; 3D LV EDVi, left ventricular end-diastolic volume index (3D echo); LA ESAi, left atrial end-systolic area index; 3D LV SVi, left ventricular stroke volume index (3D echo); MV E/A, mitral valve early-to-late filling velocity ratio; BPsys peak, systolic blood pressure at peak exercise; BP, blood pressure; PWV, pulse wave velocity; Ao. imp. Index, aortic impedance index; Br. PWV left m., brachial pulse wave velocity left measurement; Br. PWV right m., brachial pulse wave velocity right measurement; FRC, functional residual capacity; P0.1, airway occlusion pressure at 0.1 s; P0.1/Pimax, ratio of airway occlusion pressure to maximal inspiratory pressure (respiratory drive index); AaDO_₂_(peak), alveolar-arterial oxygen difference at peak exercise; DLCO/VA, diffusion capacity per unit alveolar volume (transfer coefficient/Krogh factor); ΔpCO_₂_, carbon dioxide partial pressure difference indicating high V/Q mismatch; RA ESAi, right atrial end-systolic area index; S’, peak systolic velocity of the lateral tricuspid annulus; SVi, stroke volume index; CVP, central venous pressure; 3D RV SVi, right ventricular stroke volume index (3D echocardiography); Vv hep vel D, hepatic vein diastolic blood flow velocity; RV EDAi, right ventricular end-diastolic area index; RA strain contr, right atrial contractile strain; Vv hep vel S, hepatic vein systolic blood flow velocity; 3D LAVi, left atrial volume index (3D echocardiography); LA strain cond, left atrial conduit strain; LA ESAi, left atrial end-systolic area index; 3D LV EF, left ventricular ejection fraction (3D echocardiography); LAVi, left atrial volume index; LV EF, left ventricular ejection fraction; E/E’, ratio of early mitral inflow velocity to mitral annular early diastolic velocity; 3D LV SVi, left ventricular stroke volume index (3D echocardiography); 3D CI, cardiac index (3D echocardiography); 3D LV ESVi, left ventricular end-systolic volume index (3D echocardiography); LA strain reserve, left atrial reservoir strain; BPsys_peak, systolic blood pressure at peak exercise; Aortic AP, aortic augmentation pressure; AIX@75%, augmentation index adjusted to a heart rate of 75 bpm; BP, blood pressure; PWV, pulse wave velocity; VO_₂_AT, oxygen uptake at the anaerobic threshold.

We found distinct patterns of parameter associations with dyspnoea across subgroups (Fig. 3b, Supplementary Fig. 1, 2). Group 1 patients showed the strongest association between high dyspnoea and left heart parameters that indicate low LV ventricular function (stroke volume and diastolic function) and were associated with impaired systemic vascular function (high BPsys(peak) and PWV), as well as signs of bronchial limitation ((ex-)smokers, sRaw tot; Fig. 3c). Groups 2 showed a weak association between high dyspnoea and BGA and RV parameters characteristic for hyperventilation and low RV function, but with simultaneously high CI as determined by 3D echocardiography (Fig. 3c). Group 3 dyspnoea was associated with pulmonary and systemic vascular parameters, in particular parameters of hyperventilation (low arterial pCO_2_ and high pH at rest, as well as low PetCO_2_ and arterial pCO_2_ during exercise), V/Q mismatch and low blood pressure (Fig. 3c).

### Subgroup-specific associations of peakVO_2_ with gas exchange and cardiac function or aerobic capacity

In Group 1, high peakVO_2_ was associated with high cardiopulmonary parameters indicating physiological limitations of exercise (Fig. 4a, b, Supplementary Fig. 3d, f). In contrast, Groups 2 and 3 patients were mainly limited by peripheral parameters (VO_2_AT) and blood pressure (Fig. 3b, Supplementary Fig. 4).

**Fig. 4.**
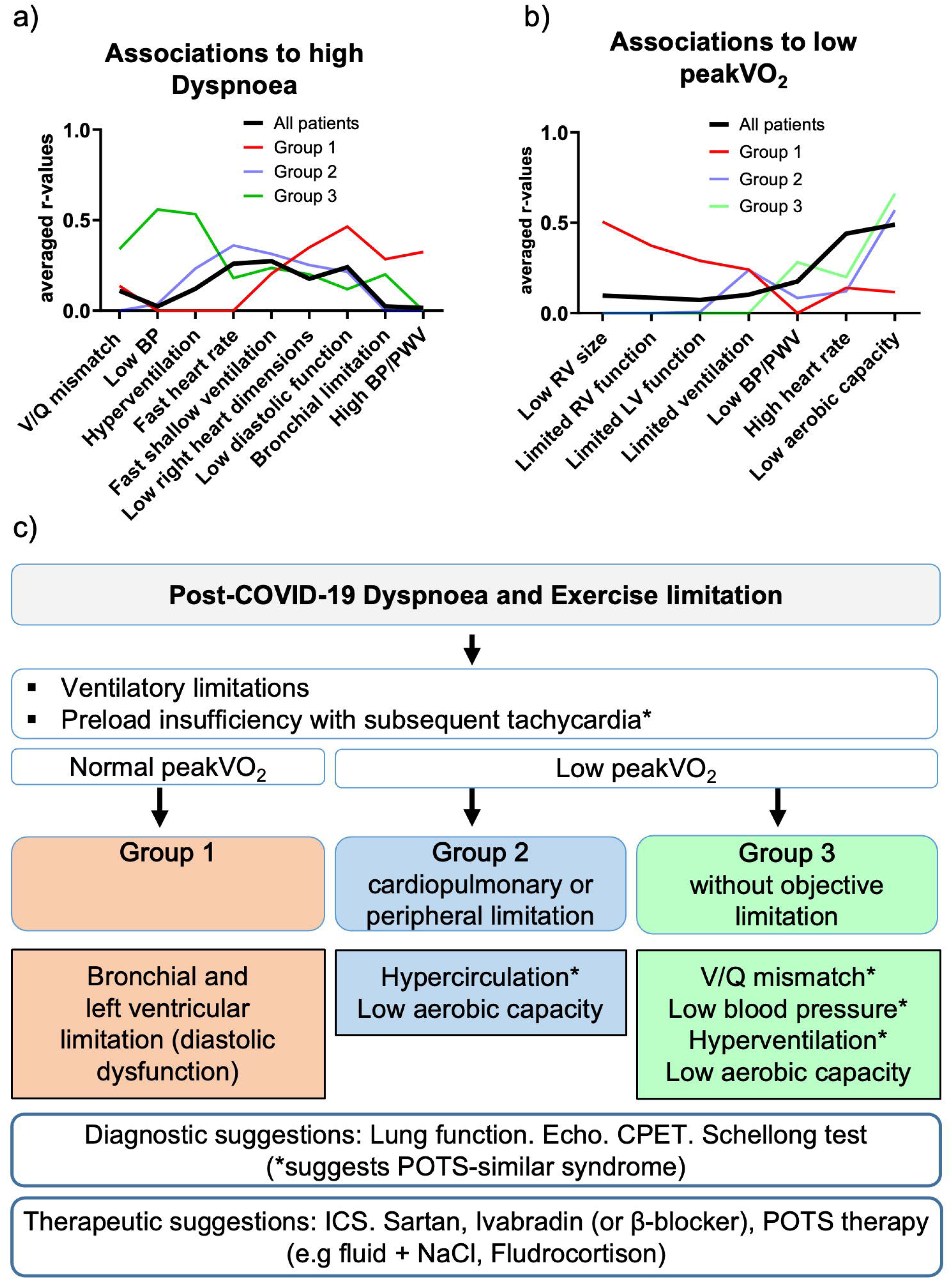
Proposed pathomechanisms for dyspnoea and exercise limitation. Associations between **a)** high dyspnoea and **b)** low peakVO_₂_ and distinct pathophysiological patterns. Fisher’s r-to-z transformation was applied to each correlation coefficient (r-value) obtained from Figures 3 or 4 (parameters with correlation coefficients <–0.4 or >0.4). Sign-corrected z-values corresponding to parameters within predefined pathophysiological categories were averaged and re-transformed to r-values. The y-axis represents the averaged correlation (r) of dyspnoea (panel a) or peak VO_₂_ (panel b) with the specific pathologies shown on the x-axis. Stronger associations indicate that a group of related parameters (representing a specific pathology) collectively contribute to symptom severity or exercise limitation. Patient subgroups are displayed separately: Group 1: normal peakVO_₂_ (% predicted); Group 2: reduced peakVO_₂_ with objective cardiopulmonary limitation on CPET; Group 3: reduced peak VO_₂_ without objective limitation on CPET. Definition of pathophysiological categories: a) V/Q mismatch: ΔpO_₂_, ΔpCO_₂_, AaDO_₂_ (baseline), VLJD/VLJT (baseline), DLCO/VA, PetCO_₂_(AT); Low blood pressure (BP): BPsys.; systolic and mean arterial pressure; Hyperventilation: pH, arterial pCO_₂_ (rest and peak), BF(peak), PetCO_₂_(AT); Fast, shallow ventilation: BF(peak), VC, FEV_₁_; Low right heart dimensions: EDVi, RA ESAi; Low diastolic function: LA strain (reservoir, contractile), MV E/A ratio; Bronchial limitation: (Ex)smoking status, RV%TLC, sRaw tot; High blood pressure (BP)/pulse wave velocity (PWV): BPsys(peak), brachial and aortic BP, brachial PWV (left) b) Low right ventricular (RV) size: RA ESAi, RV EDAi; Limited RV function: S’, SVi, 3D RV SVi, RA strain (contractile), Limited LV function: LA strain (conduit), 3D LV EF, LV EF, E/E’, 3D LV SVi, 3D CI, LA strain (reservoir), O_₂_ pulse (rest); Limited ventilation: VLJE, FRC, P0.1, P0.1/Pimax, RV%TLC, BF(rest); Low BP/PWV: Aortic AP, AIX@75%, aortic PWV, brachial BP, brachial PWV (left and right), BPsys(peak); Low aerobic capacity: VO_₂_@AT, lactate/VO_₂_peak ratio. c) Proposed mechanisms of dyspnoea and exercise limitation in post-COVID-19 patients across clinical subgroups. Illustrated are the hypothesised pathophysiological mechanisms contributing to exertional dyspnoea in all post-COVID patients and in patients stratified by cardiopulmonary exercise test (CPET)-defined subgroups. In the overall cohort, the sensation of dyspnoea is related to ventilatory parameters, in particular, high breathing frequency during exercise and reduced lung volumes. Importantly, preload insufficiency appears to be a key contributor to symptom burden. In Group 1 (patients with normal peak VO_₂_), bronchial obstruction and LV diastolic dysfunction – potentially linked to endothelial impairment – may exacerbate dyspnoea. In contrast, Groups 2 and 3 (patients with reduced peak VO_₂_) demonstrate distinct profiles. Group 2: objective cardiopulmonary and/or peripheral limitations, characterised by hypercirculation and tachycardia, and impaired aerobic capacity. Group 3: absence of objective CPET abnormalities, but evidence of hyperventilation, low blood pressure and reduced aerobic capacity. *These symptoms are related to postural-tachycardia syndrome (POTS) which may be the common denominator in post-COVID syndrome. However, the results also highlight the heterogeneity of dyspnoea mechanisms in post-COVID patients and underscore the importance of individualised phenotyping for targeted diagnostic and therapeutic strategies.

### Laboratory findings

Laboratory testing did not, on average, reveal pathological values (Supplementary Table S9). The most prominent associations were between low sodium levels and dyspnoea, and between low vitamin D, calcium and CD8+ T cells with peakVO_2_ in Group 3 patients.

## Discussion

This study provides a comprehensive analysis of the pathophysiological mechanisms underlying post-COVID dyspnoea and exercise limitations by systematically investigating associations of symptoms with cardiopulmonary and metabolic findings.

We identified for the first time reduced right heart dimensions in post-COVID patients as a potential diagnostic parameter for pre-load insufficiency underlying post-COVID symptoms. Moreover, several non-pathological findings were associated with dyspnoea and/or exercise limitation, indicating that even if in the normal range, a combination of parameters may be characteristic for post-COVID patients and contribute to their symptoms. Across all patients, high dyspnoea was linked to high heart rate, fast breathing during exercise and low lung volumes, which are all signs of preload insufficiency and low diastolic function (Fig. 5a), while low peakVO_2_ was associated with high heart rate and low aerobic capacity (Fig. 5b). Importantly, we identified ventilatory limitations and preload insufficiency as the main independent contributors to dyspnoea specifically in post-COVID patients, as well as LV function and aerobic capacity predominantly limiting exercise capacity, but these were not specifically associated with post-COVID patients. In Group 1 patients with normal peakVO_2_, bronchial and left ventricular diastolic function (associated with systemic vascular dysfunction) may specifically affect the sensation of dyspnoea and exercise capacity (“Co-morbidity phenotype”), while Groups 2 and 3 patients may be limited by low aerobic capacity and either hypercirculation/tachycardia (“Tachycardic-peripheral phenotype”, Group 2), or V/Q mismatch, low blood pressure and hyperventilation resembling symptoms similar to POTS (“POTS-like phenotype”, Group 3) (Fig. 4c).

Dyspnoea is a subjective symptom that is caused by a mismatch of the central ventilatory drive to sensory feedback from (cardio)-pulmonary mechanoreceptors [30]. Thus, patients with subtle bronchial obstruction and hyperventilation may feel severely dyspnoeic despite subclinical findings. Smokers were described as being susceptible to post-COVID dyspnoea, as seen in our Group 1 patients [31]. Group 3 patients showed pronounced hyperventilation, and maximal breathing frequency in CPET correlated with high dyspnoea in the total cohort. Accordingly, dyspnoea was associated with low lung volume, which supports the concept of mechanical and central regulatory limitations in post-COVID patients, leading to relatively fast and shallow respiration. A low ventilated alveolar volume has been previously described in post-COVID and termed a “complex restriction” associated with low DLCO at normal Krogh coefficient (DLCO/VA) in lung function [32, 33]. Decreased respiratory capacity (P0.1/Pimax) may contribute to ventilatory limitations [34]. In contrast, we did not find alterations in gas exchange or hypoxaemia during exercise, although some patients exhibited signs of shunting at rest, as previously described [35]. Along this line, platypnoea-orthodeoxia syndrome, characterised by positional dyspnoea and arterial desaturation when upright, represents a unique complication increasingly recognised in post-COVID syndrome [36]. Therefore, shunting and V/Q mismatch may be compensated for during exercise and when upright position, e.g. by high breathing frequency, hypercirculation or peripheral left-to-right shunting.

Importantly, the observed low PetCO_2_ values (AT) in post-COVID patients suggest a combination of hyperventilation, dead space ventilation, and impaired heart function, leading to V/Q mismatch [37]. Thus, besides ventilatory alterations, circulatory impairment may contribute to the sensation of dyspnoea. In contrast to previous studies suggesting the persistence of RV dysfunction in post-COVID patients [15], we found that low right heart volumes correlated with dyspnoea in all patients. Thus, these patients may at least partially suffer from preload insufficiency characterised by low right atrial pressures, low stroke volume (and compensatory tachycardia), V/Q mismatch and low DLCO, and was identified as a significant contributor to unexplained dyspnoea [13, 27]. Interestingly, preload insufficiency is also a factor in POTS, which is a common finding in post-COVID patients [38]. POTS-associated low systemic blood pressure and activation of systemic (and potentially also atrial) [39] baroreceptors may play a role in the sensation of dyspnoea and hyperventilation [14]. Independently from right heart filling insufficiency, left ventricular dysfunction may contribute to dyspnoea in Group 1 patients. In this regard, left ventricular dysfunction has been described in several studies as part of the long-term sequelae of COVID-19 [16, 40, 41], and may contribute to dyspnoea and fast, shallow breathing triggered by pulmonary juxtacapillary receptors in response to increased pressure in the capillaries [14].

In addition, it has been speculated that increased lactate production due to compromised aerobic capacity promotes hyperventilation [42]. This concept is supported by our findings of the association between dyspnoea with lactate levels in Group 3 patients and limitations in aerobic capacity in Groups 2 and 3 patients. It has been speculated, and shown for post-exertional malaise, that muscle impairment contributes to low oxygen uptake in these patients [43]. However, we cannot distinguish if decreased aerobic capacity and low oxygen uptake are due to deconditioning, perfusion disturbances or primary muscle/mitochondrial abnormalities. At least in Group 2 patients, we found an association between high cardiac output and dyspnoea, suggesting – similar to Singh et al. [10] that in these patients, peripheral oxygen extraction is limited.

Our findings strongly support the concept of dysregulation of the autonomic nervous system as an integrative cause of dyspnoea and exercise limitation in post-COVID patients characterised by preload insufficiency, tachycardia and low blood pressure [1], as previously described for other viral infections, including influenza and EBV [44]. This autonomous dysfunction may simply be caused by viral damage of the afferent pulmonary stretch sensors in the bronchial epithelium that are innervated by the vagus nerve [31] and subsequently irritated autonomous lung-brain circuits that regulate ventilation. However, systemic and in particular glial inflammation, as suggested in post-COVID syndrome [14], could also result in overactivation of the sympathetic nervous system [45].

### Limitations of the study

This study was designed as a cross-sectional multicentre trial. Limited availability of investigational methods at different study sites led to a high number of missing values of some parameters (e.g. 3D echocardiography and non-invasive determination of systemic vascular function). Previous studies stress that these trials must be well-powered and designed to accommodate the heterogeneity of long COVID presentations [1, 46]. Our study, while cross-sectional and limited in the number of participants, highlights distinct mechanisms contributing to dyspnoea and impaired exercise capacity, and thus deep-phenotyping is a suitable approach to generate hypotheses that need to be proven in further cohorts. Future research should include long-term follow-up to monitor disease trajectories and identify biomarkers for early intervention.

In summary, we identified different phenotypes of post-COVID patients in which the following factors may contribute to dyspnoea and exercise limitation to different degrees: i) circulatory limitations due to preload insufficiency and tachycardia with relatively high (Group 1), normal (Group 2) or low blood pressure (Group 3); ii) co-morbidities, such as bronchial and cardiovascular limitations (Group 1) and low LV diastolic function (all groups); iii) ventilatory limitations associated with relatively fast, shallow breathing (all groups); and iv) limited aerobic capacity (Groups 2 and 3). Addressing autonomous dysfunction and graded exercise programs may be promising therapeutic approaches. Thorough clinical work-up and classification of post-COVID patients is essential for the treatment and design of future interventional trials.

## Data Availability

All data produced in the present work are contained in the manuscript.

## Acknowledgements

The assistance of M. Wulf, D. Grund, Y. Li, M. Müller-P., Jörg C. Wildberg, E. Zessin and M. Pileckaite was highly appreciated.

## Footnotes

Support Statement: The study was funded by the Federal Ministry of Education and Research (BMBF) (AZ: 01EP2102A).

G. Oruqaj, K. Lo, Z. Rako, N. Kremer,S. Yildiz, M. Hecker, M. Kaya, S. Kuhnert, J. Wilhelm, Weissmann, H.-A. Ghofrani, F. Grimminger, W. Seeger, K. Tello and N. Sommer were supported by the Excellence Cluster Cardio-Pulmonary Institute (CPI).

K. Lo was funded by the Justus Liebig University (JLU) CAREER Programm, number GU 405/14-1. C. Tabeling received honoraria for lectures and advisory from AstraZeneca, Berlin-Chemie, GlaxoSmithKline, and for non-financial support from AstraZeneca and GlaxoSmithKline.

S. Herold was supported by the German Research Foundation grant KFO309 (reference number 284237345, projects P2, P8); Hessen State Ministry of Higher Education, Research and the Arts (HMWK, Pandemic Network Hessen and Landes-Offensive zur Entwicklung Wissenschaftlich-oekonomischer Exzellenz, LOEWE, Foerderlinie 4a project ID III L7-519/05.00.002 and CoroPan P2); the German Center for Lung Research (DZL, reference number 82DZL005B1 and 82DZLT85C1), the German Center for Infection Research (DZIF); the Excellence Cluster Cardio-Pulmonary System/Cardio-Pulmonary Institute (EXC 2026, reference number 390649896); and the Network of University Medicine (NUM, Collaborative Immunity Platform of the NUM (COVIM), NUM Study Network Infections, German National Pandemic Cohort Network (NAPKON), reference number 01KX2121); VolkswagenStiftung (project Swarm Learning).

N. Sommer was supported by the German Research Foundation grant KFO309 (reference number 284237345, project P10), the Excellence Cluster Cardio-Pulmonary System/Cardio-Pulmonary Institute (EXC 2026, reference number 390649896) and the Network of University Medicine (NUM, Collaborative Immunity Platform of the NUM, COVIM).

## References

1. Davis HE, McCorkell L, Vogel JM, Topol EJ. Long COVID: major findings, mechanisms and recommendations. Nat Rev Microbiol. 2023;21(3):133–46.

2. Ely EW, Brown LM, Fineberg HV. Long Covid Defined. N Engl J Med. 2024;391(18):1746–53.

3. Gogoll C, Peters EMJ, Köllner V, Koczulla AR. [Not Available]. Pflege Z. 2023;76(6):30–3.

4. Yu JZ, Granberg T, Shams R, Petersson S, Sköld M, Nyrén S, et al. Lung perfusion disturbances in nonhospitalized post-COVID with dyspnea-A magnetic resonance imaging feasibility study. J Intern Med. 2022;292(6):941–56.

5. Munker D, Veit T, Barton J, Mertsch P, Mümmler C, Osterman A, et al. Pulmonary function impairment of asymptomatic and persistently symptomatic patients 4 months after COVID-19 according to disease severity. Infection. 2022;50(1):157–68.

6. Azcue N, Del Pino R, Acera M, Fernández-Valle T, Ayo-Mentxakatorre N, Pérez-Concha T, et al. Dysautonomia and small fiber neuropathy in post-COVID condition and Chronic Fatigue Syndrome. J Transl Med. 2023;21(1):814.

7. Kedor C, Freitag H, Meyer-Arndt L, Wittke K, Hanitsch LG, Zoller T, et al. Author Correction: A prospective observational study of post-COVID-19 chronic fatigue syndrome following the first pandemic wave in Germany and biomarkers associated with symptom severity. Nat Commun. 13. England2022. p. 6009.

8. Legler F, Meyer-Arndt L, Mödl L, Kedor C, Freitag H, Stein E, et al. Long-term symptom severity and clinical biomarkers in post-COVID-19/chronic fatigue syndrome: results from a prospective observational cohort. EClinicalMedicine. 2023;63:102146.

9. Kuodi P, Gorelik Y, Gausi B, Bernstine T, Edelstein M. Characterization of post-COVID syndromes by symptom cluster and time period up to 12 months post-infection: A systematic review and meta-analysis. Int J Infect Dis. 2023;134:1–7.

10. Singh I, Joseph P, Heerdt PM, Cullinan M, Lutchmansingh DD, Gulati M, et al. Persistent Exertional Intolerance After COVID-19: Insights From Invasive Cardiopulmonary Exercise Testing. Chest. 2022;161(1):54–63.

11. Atchison CJ, Davies B, Cooper E, Lound A, Whitaker M, Hampshire A, et al. Long-term health impacts of COVID-19 among 242,712 adults in England. Nat Commun. 2023;14(1):6588.

12. Rinaldo RF, Mondoni M, Parazzini EM, Pitari F, Brambilla E, Luraschi S, et al. Deconditioning as main mechanism of impaired exercise response in COVID-19 survivors. Eur Respir J. 58. England2021.

13. Tooba R, Mayuga KA, Wilson R, Tonelli AR. Dyspnea in Chronic Low Ventricular Preload States. Ann Am Thorac Soc. 2021;18(4):573–81.

14. Daly ML. Hypoventilation. In: Deborah C. Silverstein KH, editor. Small Animal Critical Care Medicine (Second Edition). 2 ed2015. p. 86-92.

15. Pagnesi M, Baldetti L, Beneduce A, Calvo F, Gramegna M, Pazzanese V, et al. Pulmonary hypertension and right ventricular involvement in hospitalised patients with COVID-19. Heart. 2020;106(17):1324–31.

16. Puntmann VO, Martin S, Shchendrygina A, Hoffmann J, Ka MM, Giokoglu E, et al. Long-term cardiac pathology in individuals with mild initial COVID-19 illness. Nat Med. 2022;28(10):2117–23.

17. Haffke M, Freitag H, Rudolf G, Seifert M, Doehner W, Scherbakov N, et al. Endothelial dysfunction and altered endothelial biomarkers in patients with post-COVID-19 syndrome and chronic fatigue syndrome (ME/CFS). J Transl Med. 2022;20(1):138.

18. Thomsen LP, Karbing DS, Smith BW, Murley D, Weinreich UM, Kjærgaard S, et al. Clinical refinement of the automatic lung parameter estimator (ALPE). J Clin Monit Comput. 2013;27(3):341–50.

19. Balady GJ, Arena R, Sietsema K, Myers J, Coke L, Fletcher GF, et al. Clinician’s Guide to Cardiopulmonary Exercise Testing in Adults. Circulation. 2010;122(2):191–225.

20. Rako ZA, Kremer N, Yogeswaran A, Richter MJ, Tello K. Adaptive versus maladaptive right ventricular remodelling. ESC Heart Fail. 2023;10(2):762–75.

21. Bauer P, Kraushaar L, Most A, Hölscher S, Tajmiri-Gondai S, Dörr O, et al. Impact of Vascular Function on Maximum Power Output in Elite Handball Athletes. Res Q Exerc Sport. 2019;90(4):600–8.

22. Rees SE, Kjaergaard S, Perthorgaard P, Malczynski J, Toft E, Andreassen S. The automatic lung parameter estimator (ALPE) system: non-invasive estimation of pulmonary gas exchange parameters in 10-15 minutes. J Clin Monit Comput. 2002;17(1):43–52.

23. Vonk MC, Sander MH, van den Hoogen FH, van Riel PL, Verheugt FW, van Dijk AP. Right ventricle Tei-index: a tool to increase the accuracy of non-invasive detection of pulmonary arterial hypertension in connective tissue diseases. Eur J Echocardiogr. 2007;8(5):317–21.

24. van der Zwaan HB, Geleijnse ML, Soliman OI, McGhie JS, Wiegers-Groeneweg EJ, Helbing WA, et al. Test-retest variability of volumetric right ventricular measurements using real-time three-dimensional echocardiography. J Am Soc Echocardiogr. 2011;24(6):671–9.

25. Krittanawong C, Maitra NS, Hassan Virk HU, Farrell A, Hamzeh I, Arya B, et al. Normal Ranges of Right Atrial Strain: A Systematic Review and Meta-Analysis. JACC Cardiovasc Imaging. 2023;16(3):282–94.

26. Oldham WM, Loscalzo J. Quantification of 2-Hydroxyglutarate Enantiomers by Liquid Chromatography-mass Spectrometry. Bio Protoc. 2016;6(16).

27. Oldham WM, Lewis GD, Opotowsky AR, Waxman AB, Systrom DM. Unexplained exertional dyspnea caused by low ventricular filling pressures: results from clinical invasive cardiopulmonary exercise testing. Pulm Circ. 2016;6(1):55–62.

28. Karunanithi Z, Andersen MJ, Mellemkjær S, Alstrup M, Waziri F, Skibsted Clemmensen T, et al. Elevated Left and Right Atrial Pressures Long-Term After Atrial Septal Defect Correction: An Invasive Exercise Hemodynamic Study. J Am Heart Assoc. 2021;10(14):e020692.

29. Schnell F, Claessen G, La Gerche A, Claus P, Bogaert J, Delcroix M, et al. Atrial volume and function during exercise in health and disease. J Cardiovasc Magn Reson. 2017;19(1):104.

30. Sharma S, Hashmi MF, Badireddy M. Dyspnea on Exertion. StatPearls. Treasure Island (FL): StatPearls Publishing Copyright © 2025, StatPearls Publishing LLC.; 2025.

31. Subramanian A, Nirantharakumar K, Hughes S, Myles P, Williams T, Gokhale KM, et al. Symptoms and risk factors for long COVID in non-hospitalized adults. Nat Med. 2022;28(8):1706–14.

32. Steinbeis F, Kedor C, Meyer HJ, Thibeault C, Mittermaier M, Knape P, et al. A new phenotype of patients with post-COVID-19 condition is characterised by a pattern of complex ventilatory dysfunction, neuromuscular disturbance and fatigue symptoms. ERJ Open Res. 2024;10(5).

33. Steinbeis F, Thibeault C, Doellinger F, Ring RM, Mittermaier M, Ruwwe-Glösenkamp C, et al. Severity of respiratory failure and computed chest tomography in acute COVID-19 correlates with pulmonary function and respiratory symptoms after infection with SARS-CoV-2: An observational longitudinal study over 12 months. Respir Med. 2022;191:106709.

34. Hennigs JK, Huwe M, Hennigs A, Oqueka T, Simon M, Harbaum L, et al. Respiratory muscle dysfunction in long-COVID patients. Infection. 2022;50(5):1391–7.

35. Harbut P, Prisk GK, Lindwall R, Hamzei S, Palmgren J, Farrow CE, et al. Intrapulmonary shunt and alveolar dead space in a cohort of patients with acute COVID-19 pneumonitis and early recovery. Eur Respir J. 2023;61(1).

36. Saeed S, Hoxha B, Rajani R, Mohamed Ali A, Lehmann S. Association between Covid-19 infection and platypnea-orthodeoxia syndrome. Ann Med Surg (Lond). 2023;85(11):5813–5.

37. Myers J, Gujja P, Neelagaru S, Hsu L, Vittorio T, Jackson-Nelson T, et al. End-tidal CO2 pressure and cardiac performance during exercise in heart failure. Med Sci Sports Exerc. 2009;41(1):19–25.

38. Mallick D, Goyal L, Chourasia P, Zapata MR, Yashi K, Surani S. COVID-19 Induced Postural Orthostatic Tachycardia Syndrome (POTS): A Review. Cureus. 2023;15(3):e36955.

39. Fukushi I, Okada Y. Mechanism of dyspnea sensation: A comprehensive review for better practice of pulmonary rehabilitation. Journal of Rehabilitation Neurosciences. 2019;19:22–32.

40. Ardahanli I, Akhan O, Sahin E, Akgun O, Gurbanov R. Myocardial performance index increases at long-term follow-up in patients with mild to moderate COVID-19. Echocardiography. 2022;39(4):620–5.

41. Puntmann VO, Nagel E. Toward Better Understanding of Cardiac Involvement Post COVID. JACC Cardiovasc Imaging. 2023;16(5):625–7.

42. de Boer E, Petrache I, Goldstein NM, Olin JT, Keith RC, Modena B, et al. Decreased Fatty Acid Oxidation and Altered Lactate Production during Exercise in Patients with Post-acute COVID-19 Syndrome. Am J Respir Crit Care Med. 2022;205(1):126–9.

43. Appelman B, Charlton BT, Goulding RP, Kerkhoff TJ, Breedveld EA, Noort W, et al. Muscle abnormalities worsen after post-exertional malaise in long COVID. Nat Commun. 2024;15(1):17.

44. Levine KS, Leonard HL, Blauwendraat C, Iwaki H, Johnson N, Bandres-Ciga S, et al. Virus exposure and neurodegenerative disease risk across national biobanks. Neuron. 2023;111(7):1086–93.e2.

45. Marina N, Teschemacher AG, Kasparov S, Gourine AV. Glia, sympathetic activity and cardiovascular disease. Exp Physiol. 2016;101(5):565–76.

46. Al-Aly Z, Bowe B, Xie Y. Long COVID after breakthrough SARS-CoV-2 infection. Nat Med. 2022;28(7):1461–7.

