## Supplemental Table for "Preload insufficiency as common denominator of exertional dyspnoea in distinct post-COVID phenotypes"

| | All patients | | | | Normal peakVO2 (Group 1) | | | | Low peakVO2, maximal effort (Group 2) | | | | Low peakVO2, submaximal effort (Group 3) | | | | Control | | | |
|---|---|---|---|---|---|---|---|---|---|---|---|---|---|---|---|---|---|---|---|---|
| | **Median/ ratio** | **0.25 Quant.** | **0.75 Quant.** | ***n*** | **Median/ ratio** | **0.25 Quant.** | **0.75 Quant.** | ***n*** | **Median/ ratio** | **0.25 Quant.** | **0.75 Quant.** | ***n*** | **Median/ ratio** | **0.25 Quant.** | **0.75 Quant.** | ***n*** | **Median/ ratio** | **0.25 Quant.** | **0.75 Quant.** | ***n*** |
| **Days after infection** | **323** | 628 | 192 | *86* | **322** | 585 | 240 | *27* | **329** | 630 | 173 | *30* | **307** | 751 | 165 | *18* | **448** | 631 | 250 | *12* |
| **Age** | **44** | 35 | 57 | *86* | **54** | 44 | 59 | *27* | **36** | 29 | 48 | *30* | **44** | 35 | 59 | *18* | **37** | 26 | 47 | *12* |
| **Gender (f/m)** | **58/28** | | | *86* | **24/3** | | | *27* | **16/14** | | | *30* | **13/5** | | | *18* | **5/7** | | | *12* |
| **BMI (kg/m²)** | **26** | 23 | 29 | *86* | **27** | 23 | 28 | *27* | **27** | 23 | 31 | *30* | **25** | 23 | 28 | *18* | **23** | 23 | 25 | *12* |
| **N° of symptoms during acute infection** | **7** | 4 | 10 | *86* | **5** | 3 | 9 | *27* | **9** | 5 | 10 | *30* | **7** | 6 | 9 | *18* | **0** | 0 | 0 | *12* |
| **Co-Morbidities** | **1** | 0 | 2 | *86* | **1** | 0 | 2 | *27* | **1** | 0 | 2 | *30* | **1** | 0 | 2 | *18* | **1** | 0 | 1 | *12* |
| **(Ex)smoking (years)** | **0** | 0 | 10 | *86* | **4** | 0 | 10 | *27* | **0** | 0 | 0 | *30* | **4** | 0 | 22 | *18* | **0** | 0 | 6 | *12* |
| **Exercise (min/week)** | **90** | 30 | 139 | *82* | **90** | 60 | 128 | *27* | **85** | 19 | 180 | *30* | **75** | 34 | 120 | *18* | **220** | 90 | 360 | *12* |
| **Exercise pre-inf.** | **225** | 135 | 360 | *85* | **195** | 120 | 338 | *27* | **285** | 141 | 420 | *30* | **195** | 158 | 240 | *18* | **245** | 83 | 360 | *12* |
| **Dyspnoea index pre-inf.** | **1** | 0 | 1 | *86* | **1** | 0 | 1 | *27* | **1** | 0 | 1 | *30* | **1** | 0 | 1 | *18* | **1** | 0 | 1 | *12* |
| **ME/CFS** | **50** | 40 | 70 | *43* | **65** | 43 | 80 | *18* | **40** | 40 | 58 | *14* | **50** | 43 | 50 | *6* | **100** | 98 | 100 | *12* |
| **Pain*** | **10** | 4 | 15 | *86* | **7** | 2 | 9 | *27* | **12** | 8 | 15 | *30* | **11** | 3 | 15 | *18* | **0** | 0 | 1 | *12* |
| **CFS Score*** | **10** | 8 | 12 | *86* | **9** | 7 | 12 | *27* | **11** | 8 | 12 | *30* | **11** | 8 | 12 | *18* | **1** | 1 | 2 | *12* |
| **Sleep Score** | **-1** | -2 | 0 | *86* | **-2** | -2 | -1 | *27* | **-1** | -2 | -1 | *30* | **-1** | -2 | 0 | *18* | **2** | 1 | 2 | *12* |
| **Fear** | **1** | 0 | 2 | *86* | **0** | 0 | 2 | *27* | **1** | 0 | 1 | *30* | **1** | 0 | 2 | *18* | **0** | 0 | 0 | *12* |
| **peakVO$_2$[#]** | **86** | 74 | 99 | *78* | **107** | 96 | 116 | *27* | **79** | 73 | 86 | *30* | **71** | 63 | 78 | *18* | **108** | 87 | 119 | *12* |
| **Performance (W)** | **125** | 116 | 169 | *78* | **150** | 123 | 150 | *27* | **127** | 125 | 180 | *30* | **103** | 98 | 125 | *18* | **200** | 175 | 213 | *12* |
| **SO$_2$ (%)** | **97** | 96 | 99 | *84* | **97** | 96 | 98 | *27* | **97** | 96 | 99 | *30* | **98** | 97 | 99 | *18* | **98** | 97 | 98 | *12* |
| **pH** | **7.42** | 7.40 | 7.43 | *86* | **7.42** | 7.41 | 7.43 | *27* | **7.41** | 7.40 | 7.43 | *30* | **7.42** | 7.41 | 7.44 | *18* | **7.41** | 7.40 | 7.43 | *12* |
| **art. pCO$_2$ (mmHg)** | **37** | 35 | 40 | *86* | **38** | 33 | 41 | *27* | **38** | 36 | 40 | *30* | **37** | 35 | 39 | *18* | **38** | 36 | 39 | *12* |
| **art. pO$_2$ (mmHg)** | **85** | 76 | 90 | *86* | **85** | 76 | 93 | *27* | **86** | 81 | 89 | *30* | **85** | 77 | 90 | *18* | **91** | 85 | 92 | *12* |
| **stand. Bicarbonate** | **25** | 23 | 25 | *80* | **25** | 23 | 26 | *26* | **25** | 23 | 25 | *29* | **24** | 23,5 | 25 | *15* | **25** | 24 | 25 | *12* |
| **Base excess (mmol/l)** | **0.1** | 2 | 1 | *79* | **0.8** | -2.4 | 1.2 | *26* | **-0.1** | -1.5 | 0.8 | *29* | **-0.2** | -2 | 0.6 | *15* | **0.3** | -0.7 | 0.9 | *12* |
| **CO Hb** | **1** | 0.9 | 1.2 | *71* | **1** | 0.9 | 1.1 | *23* | **1** | 0.8 | 1.2 | *26* | **1.1** | 0.9 | 1.2 | *13* | **1.4** | 1.2 | 1.6 | *4* |
| **VC (% pred.)** | **102** | 94 | 108 | *86* | **105** | 98 | 111 | *27* | **98** | 92 | 105 | *30* | **102** | 93 | 108 | *18* | **103** | 94 | 109 | *12* |
| **FEV1 (% pred.)** | **98** | 88 | 103 | *86* | **99** | 97 | 103 | *27* | **97** | 88 | 103 | *30* | **99** | 89 | 103 | *18* | **97** | 93 | 102 | *12* |
| **sRaw tot (% pred.)** | **0.90** | 0.68 | 0.99 | *86* | **0.78** | 0.71 | 0.99 | *27* | **0.78** | 0.59 | 0.90 | *30* | **0.87** | 0.70 | 1.19 | *18* | **0.94** | 0.73 | 1.13 | *12* |
| **RV%TLC** | **34** | 28 | 47 | *86* | **36** | 31 | 46 | *27* | **31** | 26 | 42 | *30* | **36** | 32 | 45 | *17* | **34** | 30 | 87 | *12* |
| **TLC (% pred.)** | **97** | 86 | 106 | *86* | **100** | 94 | 107 | *27* | **95** | 82 | 104 | *30* | **97** | 87 | 105 | *18* | **98** | 95 | 105 | *12* |
| **BF(rest)** | **17** | 14 | 19 | *68* | **14** | 12 | 19 | *23* | **17** | 15 | 20 | *28* | **17** | 15 | 19 | *14* | **20** | 18 | 23 | *12* |
| **BF(peak)** | **34** | 29 | 39 | *73* | **36** | 30 | 42 | *26* | **35** | 30 | 39 | *28* | **30** | 24 | 32 | *16* | **43** | 39 | 49 | *12* |
| **VE(rest) (l/min)** | **11** | 9 | 14 | *78* | **10** | 9 | 14 | *27* | **11** | 9 | 16 | *30* | **10** | 8 | 12 | *18* | **15** | 11 | 18 | *12* |
| **VE(peak) (l/min)*[#]** | **61** | 52 | 78 | *78* | **63** | 54 | 89 | *27* | **64** | 56 | 83 | *30* | **48** | 40 | 56 | *18* | **78** | 71 | 108 | *12* |
| **Vtex(rest) (l)** | **0.65** | 0.50 | 0.80 | *74* | **0.67** | 0.55 | 0.82 | *26* | **0.67** | 0.49 | 0.78 | *28* | **0.59** | 0.46 | 0.73 | *17* | **0.75** | 0.56 | 0.87 | *12* |
| **Vtex(peak) (l)** | **1.87** | 1.49 | 2.97 | *74* | **1.87** | 1.59 | 2.09 | *26* | **1.90** | 1.44 | 2.37 | *28* | **1.52** | 1.46 | 2.37 | *17* | **2.09** | 1.79 | 2.29 | *12* |
| **P0.1** | **0.21** | 0.16 | 0.3 | *74* | **0.21** | 0.15 | 0.32 | *23* | **0.18** | 0.15 | 0.26 | *28* | **0.25** | 0.17 | 0.31 | *16* | **0.22** | 0.16 | 0.24 | *12* |
| **Pimax (cmH$_2$O)** | **5.41** | 4.24 | 7.48 | *74* | **5.15** | 3.01 | 8.2 | *23* | **5.60** | 4.72 | 6.52 | *29* | **5.20** | 4.25 | 7.40 | *16* | **7.62** | 6.27 | 8.78 | *12* |
| **P0.1/Pimax** | **0.04** | 0.03 | 0.06 | *74* | **0.05** | 0.03 | 0.07 | *23* | **0.04** | 0.03 | 0.05 | *29* | **0.05** | 0.03 | 0.06 | *16* | **0.03** | 0.02 | 0.03 | *12* |

*Supplemental Table S1:* Demographic characteristics, symptom load, vital parameters, ventilatory parameters and inspiratory muscle measurements of all patients, patients with normal peakVO$_2$ (Group 1), patients with reduced peakVO$_2$ and maximal effort (Group 2), patients with reduced peakVO$_2$ (Group 3) and control group. CFS scores (10 out of 11), pain (10 out of a maximum of 20 points), and fear scores (1 out of 4), sleep quality (-1, on a scale from -2 for poor sleep to +2 for good sleep) and ME/CFS Bell scores (50 out of a maximum of 100). Parameters with p<0.05 are highlighted in gray. *: significant differences (tested with ttest between all patients and control group) , [#]: significant differences (tested with oneway ANOVA and posthoc for group 1, 2 and 3, significany for group 1 and 3).

*f/m: female/male, BMI: body mass index, N°: number. pre-inf.: pre-infection, ME/CFS Score: myalgic encephalomyelitis/chronis fatigue score, CFS score: clinical frailty scale score, peakVO$_2$: oxygen uptake at peak exercise. pred.: predicted, SO$_2$: oxygen saturation, art.: arterial, art. pCO$_2$: partial pressure of carbon dioxide, art. pO$_2$: partial pressure of oxygen. VC: vital capacity. FEV1: forced expiratory volume in 1 s, sRaw tot: total specific resistance, RV%TLC: residual volume in % total lung capacity, TLC: total lung capacity, CO Hb: Carboxyhaemoglobin, P0.1: airway occlusion pressure, Pimax: maximal inspiratory pressure, VE: ventilation, BF: Breath rate*

| | **All patients** | | | | **Normal peakVO2 (Group 1)** | | | | **Low peakVO2, maximal effort (Group 2)** | | | | **Low peakVO2, submaximal effort (Group 3)** | | | | **Control** | | | |
|---|---|---|---|---|---|---|---|---|---|---|---|---|---|---|---|---|---|---|---|---|
| | **Median** | **0.25 Quant.** | **0.75 Quant.** | ***n*** | **Median** | **0.25 Quant.** | **0.75 Quant.** | ***n*** | **Median** | **0.25 Quant.** | **0.75 Quant.** | ***n*** | **Median** | **0.25 Quant.** | **0.75 Quant.** | ***n*** | **Median** | **0.25 Quant.** | **0.75 Quant.** | ***n*** |
| **Shunt (%)** | **7.0** | 2.1 | 11.1 | *75* | **7.1** | 3.3 | 10.2 | *27* | **7.0** | 3.0 | 12.5 | *25* | **4.5** | 0.3 | 9.7 | *14* | **4.8** | 1.3 | 11.3 | *11* |
| **$\Delta pO_2$ (kPa/l/min)** | **2.0** | 0.8 | 3.0 | *75* | **2.0** | 0.6 | 2.5 | *27* | **2.0** | 1.0 | 2.0 | *25* | **2.5** | 1.0 | 3.0 | *14* | **1.0** | 0.0 | 2.0 | *11* |
| **$\Delta pCO_2$ (kPa/l/min)** | **0.8** | 0.1 | 1.7 | *75* | **0.9** | 0.1 | 1.7 | *27* | **0.4** | 0.1 | 1.3 | *25* | **1.3** | 0.8 | 1.9 | *14* | **0.5** | 0.1 | 1.2 | *11* |
| **DLCO (% pred.)** | **78** | 67 | 83 | *84* | **78** | 70 | 84 | *27* | **77** | 67 | 80 | *29* | **74** | 61 | 81 | *17* | **84** | 75 | 88 | *12* |
| **DLCO/VA (% pred.)** | **86** | 74 | 96 | *78* | **83** | 73 | 96 | *25* | **89** | 74 | 93 | *28* | **82** | 74 | 87 | *17* | **85** | 78 | 96 | *12* |
| **$EqCO_2$(rest)** | **33** | 30 | 36 | *76* | **33** | 30 | 37 | *27* | **33** | 30 | 36 | *30* | **32** | 31 | 33 | *16* | **31** | 29 | 32 | *12* |
| **$EqCO_2$(peak)** | **32** | 29 | 34 | *76* | **32** | 39 | 35 | *27* | **32** | 30 | 35 | *30* | **30** | 29 | 32 | *16* | **30** | 28 | 32 | *12* |
| **$EqCO_2$AT** | **28** | 27 | 31 | *74* | **29** | 27 | 30 | *27* | **28** | 27 | 30 | *29* | **29** | 27 | 32 | *15* | **25** | 23 | 27 | *12* |
| **$VE/VCO_2$** | **29** | 25 | 33 | *76* | **30** | 25 | 33 | *27* | **29** | 27 | 33 | *30* | **28** | 25 | 32 | *16* | **25** | 23 | 30 | *12* |
| **$PetCO_2$(rest) (kPa)[#]** | **4.10** | 3.78 | 4.39 | *78* | **4.10** | 3.74 | 4.36 | *27* | **4.08** | 3.74 | 4.44 | *30* | **4.15** | 3.92 | 4.20 | *18* | **4.35** | 4.01 | 4.73 | *12* |
| **$PetCO_2$AT (kPa)*** | **4.7** | 4.4 | 5.1 | *77* | **4.8** | 4.6 | 5.1 | *27* | **4.7** | 4.2 | 5.0 | *29* | **4.7** | 4.3 | 5.1 | *18* | **5.3** | 5.2 | 6.0 | *12* |
| **$PetCO_2$(peak) (AT)** | **4.5** | 4.1 | 4.9 | *78* | **4.5** | 4.2 | 5.1 | *27* | **4.3** | 3.9 | 4.7 | *30* | **4.6** | 4.2 | 5.0 | *18* | **4.7** | 4.3 | 5.2 | *12* |
| **Vd/Vt(rest) (%)** | **10** | 7 | 12 | *35* | **10** | 7 | 11 | *13* | **10** | 9 | 13 | *13* | **8** | 3 | 11 | *6* | **7** | 7 | 12 | *9* |
| **Vd/Vt(peak) (%)** | **12** | 10 | 17 | *59* | **12** | 11 | 16 | *20* | **14** | 10 | 18 | *25* | **12** | 10 | 16 | *11* | **10** | 8 | 11 | *10* |
| **art. $pO_2$(rest) (mmHg)** | **85** | 78 | 90 | *86* | **83** | 76 | 92 | *27* | **86** | 84 | 90 | *30* | **83** | 75 | 89 | *18* | **91** | 84 | 92 | *12* |
| **art. $pO_2$(peak) (mmHg)** | **91** | 84 | 95 | *80* | **88** | 83 | 94 | *27* | **93** | 89 | 97 | *30* | **89** | 81 | 91 | *18* | **88** | 83 | 99 | *12* |
| **art. $pCO_2$(rest)(mmHg)** | **38** | 35 | 40 | *86* | **38** | 34 | 41 | *27* | **38** | 36 | 40 | *30* | **37** | 35 | 39 | *18* | **37** | 35 | 39 | *12* |
| **art. $pCO_2$(peak) (mmHg)** | **36** | 33 | 38 | *78* | **36** | 34 | 38 | *26* | **34** | 32 | 37 | *30* | **36** | 33 | 38 | *18* | **36** | 34 | 38 | *12* |
| **$AaDO_2$(rest) (mmHg)** | **15** | 12 | 22 | *66* | **16** | 12 | 24 | *24* | **15** | 11 | 20 | *25* | **15** | 12 | 21 | *14* | **11** | 9 | 19 | *10* |
| **$AaDO_2$(peak) (mmHg)** | **23** | 17 | 28 | *67* | **24** | 20 | 29 | *24* | **23** | 16 | 27 | *26* | **19** | 17 | 26 | *14* | **29** | 23 | 31 | *10* |

*Supplemental Table S2:* Parameters of V/Q-Mismatch for all groups: All patients, patients with normal $peakVO_2$ (Group 1), patients with reduced $peakVO_2$ and maximal effort (Group 2), patients with reduced $peakVO_2$ (Group 3) and control group. Parameters with $p<0.05$ are highlighted in gray. *: significant differences (tested with ttest between all patients and control group) , [#]: significant differences (tested with oneway ANOVA and posthoc for group 1, 2 and 3, significany for group 1 and 3).

*$\Delta pO_2$: venous-to-arterial oxygen difference, $\Delta pCO_2$: venous-to-arterial carbon dioxide, DLCO: diffusing capacity for carbon monoxide, DLCO/VA: diffusing capacity/Alveolar volume, $EqCO_2$: ventilatory equivalent of carbon dioxide, $VE/VCO_2$: ventilation/carbon dioxide production, $PetCO_2$: end-tidal carbon dioxide tension, Vd/Vt: ratio of physiologic dead space over tidal volume, $AaDO_2$: alveolar-arterial oxygen gradient*

| | All patients | | | | Normal peak$VO_2$ (Group 1) | | | | Low peak$VO_2$, maximal effort (Group 2) | | | | Low peak$VO_2$, submaximal effort (Group 3) | | | | Control | | | |
|---|---|---|---|---|---|---|---|---|---|---|---|---|---|---|---|---|---|---|---|---|
| | **Median** | **0.25 Quant.** | **0.75 Quant.** | ***n*** | **Median** | **0.25 Quant.** | **0.75 Quant.** | ***n*** | **Median** | **0.25 Quant.** | **0.75 Quant.** | ***n*** | **Median** | **0.25 Quant.** | **0.75 Quant.** | ***n*** | **Median** | **0.25 Quant.** | **0.75 Quant.** | ***n*** |
| **sPAP (mmHg)** | **24** | 20 | 27 | *40* | **24** | 22 | 27 | *19* | **27** | 18 | 29 | *9* | **23** | 20 | 26 | *7* | **20** | 18 | 26 | *6* |
| **TAPSE (mm)** | **24** | 22 | 26 | *80* | **24** | 22 | 26 | *25* | **23** | 21 | 25 | *27* | **24** | 20 | 25 | *17* | **25** | 22 | 27 | *12* |
| **Tei-index** | **0.42** | 0.34 | 0.49 | *64* | **0.41** | 0.39 | 0.48 | *17* | **0.41** | 0.33 | 0.51 | *22* | **0.46** | 0.34 | 0.51 | *14* | **0.33** | 0.31 | 0.39 | *11* |
| **ET/AT** | **2.41** | 2.20 | 2.83 | *77* | **2.56** | 2.36 | 2.93 | *23* | **2.38** | 2.16 | 2.80 | *27* | **2.25** | 2.16 | 2.91 | *16* | **2.27** | 2.18 | 2.53 | *12* |
| **FAC (%)** | **44** | 40 | 49 | *77* | **43** | 42 | 49 | *25* | **44** | 40 | 49 | *25* | **43** | 40 | 48 | *17* | **40** | 37 | 45 | *12* |
| **S' (cm/s)** | **13** | 11 | 14 | *80* | **12** | 11 | 13 | *25* | **13** | 11 | 14 | *27* | **12** | 11 | 13 | *17* | **12** | 12 | 14 | *11* |
| **S'/RAI** | **1.91** | 1.61 | 2.13 | *79* | **1.82** | 1.46 | 2.12 | *25* | **1.94** | 1.60 | 2.28 | *27* | **1.90** | 1.70 | 2.12 | *17* | **1.51** | 1.30 | 1.82 | *11* |
| **IVCT (ms)** | **66** | 59 | 72 | *64* | **69** | 61 | 73 | *16* | **65** | 48 | 68 | *23* | **76** | 65 | 90 | *14* | **50** | 49 | 58 | *11* |
| **IVRT (ms)** | **68** | 55 | 80 | *63* | **70** | 66 | 80 | *16* | **61** | 49 | 79 | *22* | **70** | 58 | 78 | *14* | **45** | 43 | 57 | *11* |
| **E/A wave*** | **1.49** | 1.27 | 2.06 | *46* | **1.85** | 1.32 | 2.11 | *13* | **1.54** | 1.26 | 2.04 | *16* | **1.43** | 1.39 | 1.57 | *11* | **1.24** | 1.16 | 1.37 | *9* |
| **TV E/E'** | **4.84** | 4.09 | 6.16 | *42* | **4.77** | 4.10 | 5.64 | *10* | **5.75** | 4.47 | 7.50 | *17* | **4.50** | 4.08 | 5.23 | *9* | **5.22** | 4.70 | 6.47 | *9* |
| **RVEDD (mm)** | **36** | 31 | 39 | *81* | **36** | 34 | 39 | *25* | **36** | 32 | 39 | *28* | **34** | 31 | 36 | *17* | **39** | 37 | 46 | *12* |
| **RA ESAi ($cm^2/m^2$)** | **6.76** | 5.20 | 7.54 | *81* | **6.77** | 6.21 | 8.07 | *25* | **6.87** | 5.28 | 7.52 | *28* | **6.46** | 5.46 | 6.94 | *17* | **8.32** | 7.04 | 10.34 | *12* |
| **RV EDAi ($cm^2/m^2$)*** | **10** | 9 | 21 | *66* | **10** | 9 | 11 | *18* | **19** | 17 | 21 | *23* | **19** | 17 | 21 | *15* | **24** | 20 | 28 | *11* |
| **RV ESAi ($cm^2/m^2$)** | **5.64** | 5.05 | 6.70 | *67* | **5.60** | 5.00 | 6.00 | *19* | **10.50** | 8.74 | 12.35 | *23* | **11.10** | 9.69 | 11.55 | *15* | **12.80** | 11.10 | 17.20 | *11* |
| **RV wall (mm)** | **4.15** | 3.60 | 5.00 | *74* | **4.00** | 3.80 | 4.70 | *23* | **4.10** | 3.55 | 4.75 | *27* | **4.60** | 4.00 | 4.90 | *15* | **3.70** | 3.35 | 4.65 | *12* |
| **RV strain fw (%)** | **-29** | -32 | -26 | *68* | **-29** | -32 | -27 | *22* | **-31** | -35 | -25 | *26* | **-29** | -35 | -27 | *14* | **-25** | -29 | -23 | *10* |
| **RA strain reserv (%)** | **49** | 40 | 59 | *58* | **45** | 39 | 51 | *17* | **53** | 45 | 64 | *22* | **48** | 45 | 59 | *13* | **39** | 33 | 45 | *8* |
| **RA strain cond (%)** | **-33** | -39 | -25 | *59* | **-29** | -35 | -22 | *17* | **-36** | -45 | -32 | *22* | **-32** | -39 | -24 | *14* | **-31** | -35 | -23 | *8* |
| **RA strain contr (%)** | **-17** | -20 | -13 | *58* | **-15** | -20 | -12 | *17* | **-17** | -20 | -15 | *22* | **-17** | -19 | -13 | *13* | **-9** | -13 | -8 | *8* |
| **CVP (mmHg)** | **5** | 5 | 5 | *74* | **5** | 5 | 5 | *23* | **5** | 5 | 5 | *26* | **5** | 5 | 5 | *15* | **5** | 5 | 5 | *11* |
| **Vv hep vel S (cm/s)** | **42** | 30 | 61 | *46* | **43** | 35 | 58 | *13* | **39** | 30 | 72 | *17* | **42** | 32 | 61 | *11* | **35** | 28 | 43 | *10* |
| **Vv hep vel D (cm/s)** | **27** | 21 | 48 | *46* | **25** | 21 | 39 | *13* | **30** | 24 | 63 | *17* | **23** | 17 | 47 | *11* | **23** | 20 | 32 | *10* |
| **Vv hep vel A (cm/s)** | **27** | 24 | 32 | *46* | **27** | 24 | 32 | *13* | **28** | 22 | 32 | *17* | **26** | 25 | 29 | *11* | **27** | 25 | 37 | *10* |
| **3D RV EDVi ($ml/m^2$)** | **58** | 51 | 65 | *58* | **58** | 52 | 63 | *19* | **58** | 52 | 63 | *23* | **58** | 46 | 66 | *13* | **70** | 63 | 73 | *9* |
| **3D RV ESVi ($ml/m^2$)** | **26** | 21 | 29 | *58* | **24** | 21 | 27 | *19* | **27** | 23 | 29 | *23* | **27** | 19 | 31 | *13* | **33** | 28 | 36 | *9* |
| **3D RV EF (%)** | **56** | 52 | 59 | *58* | **56** | 54 | 60 | *19* | **55** | 50 | 58 | *23* | **57** | 54 | 59 | *13* | **55** | 53 | 55 | *9* |
| **3D RV SVI ($ml/m^2$)** | **32** | 28 | 36 | *57* | **34** | 30 | 36 | *19* | **31** | 28 | 34 | *23* | **29** | 24 | 37 | *13* | **36** | 36 | 39 | *9* |

*Supplemental Table S3:* Right heart parameters at rest for groups: All patients, patients with normal peak$VO_2$ (Group 1), patients with reduced peak$VO_2$ and maximal effort (Group 2), patients with reduced peak$VO_2$ (Group 3) and control group. Parameters with $p<0.05$ are highlighted in gray. *: significant differences (tested with ttest between all patients and control group).

*sPAP: systolic pulmonary artery pressure. TAPSE: tricuspid annular plane systolic excursion. Tei-index: myocardial performance index, ET/AT: ejection time/acceleration time, FAC: fractional area change, IVCT: isovolumic contraction time, IVRT: isovolumic relaxation time*, *S': systolic contraction velocity, S'/RAi: systolic contraction velocity/right atrial index, TV: tricuspid valve, RVEDD: right ventricular end-diastolic diameter*, *E/A wave: early to late diastolic transtricuspid flow velocity, E/E': ratio of the E wave velocity to myocardial movement velocity. RA: right atrium, ESAi: end-systolic area index, RV: right ventricular, CVP: central venous pressure, EDD: end-diastolic diameter, EDAi: end-diastolic area index, ESAi: end-systolic area index, EDVi: end-diastolic volume index, ESVi: end-systolic volume index, SVi: stroke volume index, EF: ejection fraction,*

| | Median/ ratio | 0.25 Quant. | 0.75 Quant. | *n* | Correlation to dysponea (r ) | Correlation to high peak $VO_2$ |
|---|---|---|---|---|---|---|
| **CPET sPAP at exercise excl. CVP** | **32.00** | 23.50 | 36.00 | *11* | -0.11 | 0.13 |
| **CPET RV Strain GLS** | **-31.95** | -34.63 | -29.00 | *22* | 0.06 | -0.03 |
| **CPET RV Strain FW** | **-34.40** | -38.48 | -29.08 | *22* | 0.03 | 0.09 |
| **CPET 3D RV EDVi** | **57.76** | 50.44 | 67.78 | *15* | -0.22 | -0.74 |
| **CPET 3D RV ESVi** | **22.30** | 19.06 | 25.63 | *15* | 0.03 | -0.62 |
| **CPET 3D RV EF** | **61.00** | 57.45 | 64.28 | *14* | -0.18 | 0.11 |
| **CPET 3D RV SVI** | **34.86** | 27.84 | 41.72 | *16* | -0.20 | -0.17 |
| **CPET LV GLS Strain** | **-27.60** | -29.40 | -25.30 | *25* | -0.14 | 0.15 |
| **CPET 3D LV EDVi** | **70.37** | 59.39 | 77.25 | *23* | -0.23 | -0.30 |
| **CPET 3D LV ESVi** | **24.83** | 21.30 | 30.48 | *23* | -0.33 | -0.32 |
| **CPET 3D LV EF** | **64** | 60 | 68 | *21* | 0.36 | 0.17 |
| **CPET 3D LV SVi** | **42.89** | 38.61 | 47.68 | *23* | -0.08 | -0.21 |
| **CPET 3D CI exercise** | **6.18** | 4.82 | 6.96 | *22* | -0.03 | -0.01 |

*Supplemental Table S4:* Echocardiography during exercise groups: All patients, patients with normal peak$VO_2$ (Group 1), patients with reduced peak$VO_2$ and maximal effort (Group 2), patients with reduced peak$VO_2$ (Group 3) and control group.

*sPAP: systolic pulmonary artery pressure, RV: right ventricular, EDVi: end-diastolic volume, ESVi: end-systolic volume, SVi: stroke volume, CPET: cardiopulmonary exercise testing, RV Strain GLS: right ventricular global longitudinal strain, RV Strain FW: right ventricular strain free wall, 3D RV EDVi: 3D Right ventricular end-diastolic volume, 3D RV ESVi: 3D Right ventricular end-systolic volume, 3D RV EF: 3D Right ventricular ejection fraction, 3D RV SVi: 3D Right ventricular stroke volume, CI: Cardiac index, 3D LV EDVi: 3D Left ventricular end-diastolic volume, 3D LV ESVi: 3D Left ventricular end-systolic volume, 3D LV SVi: 3D Left ventricular stroke volume, 3D LV EF: 3D Left ventricular ejection fraction, LV GLS Strain: left ventricular global longitudinal strain, 3D LV SVi: 3D Left ventricular stroke volume, 3D CI: 3D Cardiac index*

| | Rest | | | | Exercise | | |
|---|---|---|---|---|---|---|---|
| | Median/ ratio | 0.25 Quant. | 0.75 Quant. | | Median/ ratio | 0.25 Quant. | 0.75 Quant. |
| **HR** | **64** | 63 | 76 | | **108** | 103 | 133 |
| **RAP** | **6.0** | 4.0 | 7.5 | | **6.0** | 4.0 | 8.5 |
| **mPAP** | **15** | 14 | 17 | | **26** | 23 | 27 |
| **PAWP** | **7** | 6 | 10 | | **11** | 8 | 13 |
| **PVR** | **90** | 82 | 114 | | **97** | 75 | 108 |
| **CI TD** | **3.39** | 2.54 | 4.34 | | **5.85** | 4.66 | 7.50 |
| **$AVDO_2$** | **4.61** | 4.42 | 4.84 | | **9.32** | 8.93 | 9.65 |
| **mPAP/CO** | | | | | **1.2** | 0.83 | 1.64 |
| **PAWP/CO** | | | | | **0.42** | 0.22 | 0.72 |

*Supplemental Table S5*: Right heart catheterization before and after exercise for all patients.

*HR: heart rate, RAP: right atrial pressure*, *mPAP: mean pulmonary arterial pressure, CO: cardiac output*, *PAWP: pulmonary artery wedge pressure*, *PVR: pulmonary vascular resistance , CI TD: cardiac index thermodilution, $AVDO_2$: arterial minus jugular oxygen saturation*

| | All patients | | | | Normal peakVO2 (Group 1) | | | | Low peakVO2, maximal effort (Group 2) | | | | Low peakVO2, submaximal effort (Group 3) | | | | Control | | | |
|---|---|---|---|---|---|---|---|---|---|---|---|---|---|---|---|---|---|---|---|---|
| | **Median** | **0.25 Quant.** | **0.75 Quant.** | ***n*** | **Median** | **0.25 Quant.** | **0.75 Quant.** | ***n*** | **Median** | **0.25 Quant.** | **0.75 Quant.** | ***n*** | **Median** | **0.25 Quant.** | **0.75 Quant.** | ***n*** | **Median** | **0.25 Quant.** | **0.75 Quant.** | ***n*** |
| **LV EF (%)** | **60** | 57 | 63 | *60* | **62** | 58 | 66 | *21* | **57** | 56 | 60 | *19* | **61** | 58 | 63 | *15* | **59** | 59 | 59 | *1* |
| **MV E/A** | **1.18** | 0.94 | 1.47 | *77* | **1.18** | 1.01 | 1.45 | *25* | **1.08** | 0.90 | 1.50 | *26* | **1.17** | 0.9 | 1.60 | *17* | **1.43** | 1.21 | 1.73 | *12* |
| **E/E'** | **7.74** | 6.35 | 9.56 | *75* | **8.17** | 6.96 | 10.81 | *25* | **7.57** | 6.22 | 9.74 | *25* | **6.90** | 6.37 | 9.20 | *16* | **6.21** | 5.42 | 6.63 | *12* |
| **LV GLS (%)** | **-22** | -24 | -20 | *57* | **-23** | -24 | -20 | *18* | **-22** | -23 | -18 | *21* | **-22** | -25 | -21 | *12* | **-21** | -22 | -19 | *9* |
| **LA ESAi (cm²/m²)** | **14** | 11 | 17 | *78* | **8** | 7 | 9 | *24* | **14** | 11 | 17 | *26* | **14** | 12 | 15 | *17* | **12** | 10 | 15 | *12* |
| **LAVI (ml/m²)** | **15** | 13 | 21 | *59* | **16** | 13 | 24 | *18* | **16** | 12 | 20 | *24* | **15** | 12 | 17 | *13* | **16** | 15 | 20 | *8* |
| **EDPWD (mm)** | **7.4** | 5.9 | 9.0 | *66* | **7.2** | 5.5 | 8.7 | *16* | **6.7** | 5.6 | 8.3 | *24* | **8.4** | 6.5 | 9.1 | *15* | **8.1** | 8.0 | 9.0 | *11* |
| **EDIVS (mm)** | **6.0** | 5.2 | 6.8 | *66* | **5.7** | 5.4 | 6.6 | *16* | **5.6** | 4.3 | 6.2 | *24* | **6.0** | 5.7 | 6.9 | *15* | **6.9** | 5.1 | 7.0 | *11* |
| **LVEDD (mm)** | **45** | 42 | 49 | *66* | **45** | 40 | 49 | *16* | **46** | 42 | 50 | *24* | **43** | 42 | 46 | *15* | **46** | 41 | 48 | *11* |
| **LVESD (mm)** | **34** | 28 | 38 | *66* | **32** | 29 | 40 | *16* | **37** | 33 | 38 | *24* | **30** | 28 | 35 | *15* | **29** | 23 | 31 | *11* |
| **LA strain Reserv (%)** | **47** | 36 | 57 | *64* | **37** | 29 | 48 | *19* | **46** | 38 | 58 | *25* | **55** | 45 | 61 | *14* | **45** | 38 | 47 | *9* |
| **LA strain Cond (%)** | **-29** | -40 | -21 | *64* | **-23** | -31 | -16 | *19* | **-31** | -41 | -21 | *25* | **-38** | -44 | -26 | *14* | **-27** | -35 | -22 | *9* |
| **LA strain Kontr (%)** | **-17** | -21 | -13 | *64* | **-17** | -19 | -12 | *19* | **-17** | -21 | -13 | *25* | **-17** | -20 | -15 | *14* | **-13** | -17 | -13 | *9* |
| **3D LAVi (ml/m²)** | **25** | 20 | 31 | *46* | **29** | 25 | 34 | *14* | **21** | 19 | 26 | *19* | **24** | 20 | 30 | *10* | **29** | 25 | 31 | *8* |
| **3D LV EF (%)** | **58** | 56 | 62 | *53* | **61** | 58 | 63 | *17* | **57** | 56 | 59 | *20* | **60** | 58 | 61 | *13* | **58** | 56 | 61 | *10* |
| **3D LA EF (%)** | **66** | 60 | 71 | *46* | **63** | 55 | 70 | *14* | **68** | 61 | 69 | *19* | **68** | 62 | 71 | *10* | **61** | 57 | 62 | *10* |
| **3D LV EDVi (ml/m²)** | **64** | 56 | 74 | *53* | **63** | 56 | 74 | *17* | **65** | 57 | 71 | *20* | **64** | 51 | 77 | *13* | **72** | 57 | 83 | *10* |
| **3D LV ESVi (ml/m²)** | **27** | 22 | 32 | *53* | **24** | 22 | 29 | *17* | **28** | 23 | 32 | *20* | **28** | 21 | 33 | *13* | **32** | 24 | 34 | *10* |
| **3D LV SVi (ml/m²)** | **37** | 34 | 43 | *52* | **38** | 35 | 45 | *17* | **36** | 34 | 39 | *19* | **37** | 32 | 44 | *13* | **41** | 39 | 47 | *10* |
| **3D CI (l/min/m²)** | **2.48** | 2.31 | 3.02 | *52* | **2.41** | 2.30 | 2.87 | *17* | **2.66** | 2.42 | 3.14 | *20* | **2.46** | 2.09 | 3.04 | *12* | **3.08** | 2.70 | 3.28 | *10* |

*Supplemental Table S6:* Left heart parameters at rest for groups: All patients, patients with normal peakVO$_2$ (Group 1), patients with reduced peakVO$_2$ and maximal effort (Group 2), patients with reduced peakVO$_2$ (Group 3) and control group. Parameters with p<0.05 are highlighted in gray. *: significant differences (tested with ttest between all patients and control group).

*LV EF: left ventricular ejection fraction, MV E/A: mitral valve E-wave/A-wave ratio, E/E': left ventricular filling pressure, EDIVS: end.diastolic volume of the left heart, EDPWD: end-diastolic posterior wall diameter, LA ESAi: left atrial end systolic area, LAVI: left atrial volume index, EDPWD: , EDIVS: , LVEDD: left ventricular end-diastolic diameter, LVESD: left ventricular end-systolic diameter*, *LA strain Reserv: left atrial strain reservoir, LA strain Cond: left atrial strain conduit, LA strain Kontr: left atrial strain conduit, 3D LV EDVi: 3D Left ventricular end-diastolic volume, 3D LV ESVI: 3D Left ventricular end-systolic volume, 3D LV SVi: 3D Left ventricular stroke volume, 3D CI: 3D Cardiac index, 3D LV EF: 3D Left ventricular ejection fraction, LV GLS Strain: left ventricular global longitudinal strain*

| | All patients | | | | Normal peak$VO_2$ (Group 1) | | | | Low peak$VO_2$, maximal effort (Group 2) | | | | Low peak$VO_2$, submaximal effort (Group 3) | | | | Control | | | |
|---|---|---|---|---|---|---|---|---|---|---|---|---|---|---|---|---|---|---|---|---|
| | **Median** | **0.25 Quant.** | **0.75 Quant.** | ***n*** | **Median** | **0.25 Quant.** | **0.75 Quant.** | ***n*** | **Median** | **0.25 Quant.** | **0.75 Quant.** | ***n*** | **Median** | **0.25 Quant.** | **0.75 Quant.** | ***N*** | **Median** | **0.25 Quant.** | **0.75 Quant.** | ***n*** |
| **HF(rest) (1/min)** | **76** | 83 | 69 | *77* | **69** | 81 | 62 | *27* | **81** | 86 | 73 | *30* | **74** | 89 | 67 | *18* | **78** | 94 | 67 | *12* |
| **HF(peak) (1/min)*#** | **147** | 164.5 | 129 | *77* | **148** | 162 | 130 | *27* | **161** | 172.3 | 143.8 | *30* | **129** | 148 | 114.5 | *18* | **178** | 181 | 169 | *12* |
| **BPsys(peak) (mmHg)** | **190** | 213 | 162 | *75* | **200** | 212 | 182 | *26* | **201** | 217 | 167 | *28* | **160** | 188 | 148 | *18* | **207** | 218 | 192 | *12* |
| **BPdias(peak) (mmHg)** | **96** | 104 | 89 | *75* | **98** | 110 | 91 | *26* | **100** | 105 | 90 | *28* | **91** | 101 | 82 | *18* | **100** | 110 | 91 | *12* |
| **$O_2$ pulse(rest) (ml)** | **4** | 5 | 3 | *77* | **5** | 6 | 3 | *27* | **4** | 5 | 3 | *30* | **4** | 6 | 3 | *16* | **6** | 8 | 4 | *12* |
| **$O_2$ pulse(peak) (ml)** | **12** | 15 | 9.5 | *77* | **12** | 15 | 11 | *27* | **11.6** | 15 | 9 | *30* | **10.7** | 13.7 | 9 | *16* | **13** | 14 | 11.3 | *12* |
| **$\Delta VO_2/\Delta W$** | **21** | 17 | 31 | *57* | **26** | 21 | 33 | *20* | **20** | 15 | 32 | *22* | **18** | 16 | 20 | *12* | **19** | 15 | 24 | *0* |
| **Resistance index (%)** | **7.6** | 6.8 | 8.3 | *57* | **8.2** | 7.0 | 9.2 | *20* | **7.4** | 6.8 | 7.9 | *22* | **7.1** | 6.6 | 7.8 | *12* | **6.6** | 6.1 | 7.2 | *10* |
| **Aortic PWV (m/s)** | **203** | 169 | 234 | *57* | **201** | 163 | 226 | *20* | **216** | 177 | 235 | *22* | **187** | 161 | 219 | *12* | **175** | 130 | 252 | *10* |
| **Aortic impedance index** | **126** | 118 | 137 | *57* | **127** | 118 | 136 | *20* | **135** | 120 | 139 | *22* | **119** | 114 | 124 | *12* | **118** | 113 | 126 | *10* |
| **Brachial BP (mmHg)** | **107** | 99 | 117 | *57* | **112** | 100 | 120 | *20* | **107** | 103 | 122 | *22* | **100** | 94 | 108 | *12* | **108** | 94 | 114 | *10* |
| **Aortic BP (mmHg)** | **2** | -3 | 7 | *57* | **7** | 2 | 12 | *20* | **-2** | -5 | 4 | *22* | **0** | -3 | 4 | *12* | **3** | -2 | 5 | *10* |
| **Aortic AP (mmHg)** | **2.80** | 8.95 | 16.28 | *56* | **16.75** | 2.75 | 23.68 | *20* | **-2.20** | -13.30 | 13.00 | *21* | **-5.85** | -9.60 | 5.10 | *12* | **-0.75** | -11.33 | 15.75 | *10* |
| **AIX@75 (%)** | **1.16** | 1.11 | 1.22 | 45 | **1.19** | 1.18 | 1.16 | 13.50 | **1.14** | 1.09 | 1.19 | 20.50 | **1.21** | 1.15 | 1.25 | 9 | **1.17** | 1.14 | 1.23 | 5 |
| **Brachial ABI left measurement** | **1.17** | 1.11 | 1.23 | 46.50 | **1.19** | 1.14 | 1.22 | 14 | **1.14** | 1.07 | 1.20 | 20.50 | **1.22** | 1.18 | 1.28 | 10 | **1.20** | 1.16 | 1.23 | 9 |
| **Brachial ABI right measurement** | **9.85** | 9.20 | 11.09 | 44 | **9.80** | 9.04 | 11.40 | 13.50 | **10.45** | 9.61 | 11.25 | 19.50 | **9.75** | 9.29 | 10.40 | 9 | **8.95** | 8.66 | 10.23 | 0 |
| **Brachial PWV left measurement** | **10.53** | 9.43 | 11.39 | 45.50 | **10.05** | 9.30 | 11.08 | 14 | **11.18** | 10.14 | 12.03 | 19.50 | **9.80** | 9.35 | 10.10 | 10 | **9.70** | 8.56 | 10.16 | 10 |

*Supplemental Table S7*: Circulatory parameter and peripheral pulse wave analysis for all groups: All patients, patients with normal peak$VO_2$ (Group 1), patients with reduced peak$VO_2$ and maximal effort (Group 2), patients with reduced peak$VO_2$ (Group 3) and control group. Parameters with $p<0.05$ are highlighted in gray. *: significant differences (tested with ttest between all patients and control group), #: significant differences (tested with oneway ANOVA and posthoc for group 1, 2 and 3, significany for group 1 and 3).

*HF: heart rate, BP: blood prssure, sys: systolic, dias: diastolic, PWV: pulse wave velocity, BP: blood pressure. AIX(%): augmentation index, AIX@75: augumentation index normalized to a heart rate of 75 beats per minute, ABI: Ancle-brachial-index*

| | All patients | | | | Normal peak$VO_2$ (Group 1) | | | | Low peak$VO_2$, maximal effort (Group 2) | | | | Low peak$VO_2$, submaximal effort (Group 3) | | | | Control | | | |
|---|---|---|---|---|---|---|---|---|---|---|---|---|---|---|---|---|---|---|---|---|
| | **Median** | **0.25 Quant.** | **0.75 Quant.** | ***n*** | **Median** | **0.25 Quant.** | **0.75 Quant.** | ***n*** | **Median** | **0.25 Quant.** | **0.75 Quant.** | ***n*** | **Median** | **0.25 Quant.** | **0.75 Quant.** | ***n*** | **Median** | **0.25 Quant.** | **0.75 Quant.** | ***n*** |
| **pH** | **7.37** | 7.32 | 7.41 | *53* | **7.37** | 7.32 | 7.42 | *22* | **7.35** | 7.31 | 7.37 | *16* | **7.38** | 7.35 | 7.42 | *12* | **7.29** | 7.28 | 7.31 | *12* |
| **Lactate (mmol/l)/ peak$VO_2$ (%)** | **0.06** | 0.04 | 0.09 | *65* | **0.05** | 0.04 | 0.07 | *21* | **0.08** | 0.06 | 0.11 | *26* | **0.05** | 0.05 | 0.08 | *16* | **0.10** | 0.06 | 0.11 | *12* |
| **Lactate(rest) (mmol/l)** | **0.92** | 0.7 | 1.1 | *66* | **0.8** | 0.71 | 0.96 | *19* | **0.10** | 0.8 | 1.17 | *28* | **0.84** | 0.69 | 0.10 | *14* | **1.06** | 0.75 | 1.43 | *11* |
| **RER(rest)** | **0.82** | 0.77 | 0.93 | *76* | **0.84** | 0.76 | 0.91 | *27* | **0.83** | 0.77 | 0.10 | *30* | **0.81** | 0.76 | 0.85 | *18* | **0.84** | 0.71 | 0.91 | *12* |
| **RER(max)** | **1.11** | 1.04 | 1.18 | *76* | **1.08** | 1.05 | 1.15 | *27* | **1.18** | 1.08 | 1.26 | *30* | **1.06** | 1.00 | 1.1 | *18* | **1.17** | 1.13 | 1.22 | *12* |
| **$VO_2$AT (%max)** | **84** | 77 | 88 | *66* | **88** | 81 | 89 | *21* | **82** | 77 | 87 | *28* | **87** | 74 | 88 | *15* | **87** | 83 | 94 | *12* |
| **$VO_2$AT (%Soll)** | **67** | 58 | 82 | *77* | **86** | 68 | 97 | *27* | **67** | 60 | 69 | *30* | **57** | 38 | 69 | *18* | **87** | 81 | 103 | *12* |
| **$VO_2$(rest) (ml/min/kg)** | **0.004** | 0.003 | 0.005 | *76* | **0.004** | 0.003 | 0.005 | *27* | **0.004** | 0.004 | 0.005 | *29* | **0.004** | 0.003 | 0.005 | *18* | **0.007** | 0.006 | 0.008 | *12* |
| **$\Delta VO_2/\Delta W$** | **9.32** | 8.41 | 9.89 | *48* | **9.56** | 9.29 | 10.22 | *16* | **8.69** | 8.21 | 9.71 | *17* | **9.29** | 8.09 | 9.82 | *11* | **9.87** | 9.27 | 10.10 | *10* |

*Supplemental Table S8:* Peripheral parameters for all groups: All patients, patients with normal peak$VO_2$ (Group 1), patients with reduced peak$VO_2$ and maximal effort (Group 2), patients with reduced peak$VO_2$ (Group 3) and control group. Parameters with $p<0.05$ are highlighted in gray. *: significant differences (tested with ttest between all patients and control group) , [1]: significant differences (tested with oneway ANOVA for group 1, 2 and 3).

RER: respiratory exchange ratio *$VO_2$AT: Oxygen consumption at anaerobic threshold, $\Delta VO_2/\Delta W$: peak oxygen consumption to watt relationship*

| | All patients | | | | Normal peakVO2 (Group 1) | | | | Low peakVO2, maximal effort (Group 2) | | | | Low peakVO2, submaximal effort (Group 3) | | | | Control | | | |
|---|---|---|---|---|---|---|---|---|---|---|---|---|---|---|---|---|---|---|---|---|
| | **Median** | **0.25 Quant.** | **0.75 Quant.** | ***n*** | **Median** | **0.25 Quant.** | **0.75 Quant.** | ***n*** | **Median** | **0.25 Quant.** | **0.75 Quant.** | ***n*** | **Median** | **0.25 Quant.** | **0.75 Quant.** | ***n*** | **Median** | **0.25 Quant.** | **0.75 Quant.** | ***n*** |
| **Hb (g/l)** | **138** | 131 | 146 | *85* | **133** | 130.5 | 140 | *27* | **142** | 132.5 | 146.75 | *30* | **137** | 127 | 141 | *17* | **136** | 130.75 | 148 | *12* |
| **Hct (l/l)** | **0.408** | 0.38 | 0.43 | *83* | **0.39** | 0.38 | 0.41825 | *27* | **0.415** | 0.3825 | 0.43 | *30* | **0.4** | 0.38 | 0.42 | *17* | **0.393** | 0.3675 | 0.42225 | *12* |
| **RBC (tera/l)** | **4.6** | 4.4 | 4.883 | *84* | **4.4** | 4.3 | 4.6 | *26* | **4.75** | 4.525 | 5 | *30* | **4.6** | 4.3 | 4.8 | *17* | **4.5** | 4.15 | 4.8 | *12* |
| **Platelets (giga/l)** | **245** | 218 | 291 | *84* | **267** | 234 | 302 | *26* | **234** | 213.25 | 290.5 | *30* | **244** | 223 | 264 | *17* | **239.5** | 213.25 | 263.5 | *12* |
| **WBC (giga/l)** | **6** | 5 | 7.2 | *84* | **5.51** | 4 | 7.28 | *26* | **6** | 5 | 7.1675 | *30* | **6** | 5 | 7 | *17* | **5.9** | 5.075 | 7.025 | *12* |
| **MCV (fl)** | **87** | 85 | 90 | *83* | **89** | 85 | 91 | *26* | **86.05** | 84 | 88 | *30* | **87** | 84.1 | 90.2 | *17* | **89.3** | 87 | 91 | *12* |
| **MCH (pg)** | **29.9** | 28.85 | 30.6 | *83* | **30.2** | 29.8 | 30.9 | *26* | **29.55** | 28.7 | 30.75 | *30* | **30** | 28.8 | 30.2 | *17* | **30.95** | 30.3 | 31.875 | *12* |
| **neutrophils (giga/l)** | **3.56** | 2.52 | 4.685 | *84* | **3.41** | 2.42 | 4.47 | *26* | **3.225** | 2.7325 | 4.0175 | *30* | **3.86** | 2.76 | 5.04 | *17* | **3.615** | 2.655 | 4.635 | *12* |
| **Lymphocytes (giga/l)** | **1.74** | 1.388 | 2.09 | *84* | **1.47** | 1.28 | 2.09 | *26* | **1.84** | 1.535 | 2.0975 | *30* | **1.59** | 1.33 | 1.94 | *17* | **1.885** | 1.445 | 2.2575 | *12* |
| **Eosinophils (giga/l)** | **0.12** | 0.08 | 0.193 | *84* | **0.11** | 0.08 | 0.17 | *26* | **0.11** | 0.08 | 0.19 | *30* | **0.1** | 0.06 | 0.22 | *17* | **0.12** | 0.07 | 0.2325 | *12* |
| **Basophils( giga/l)** | **0.04** | 0.03 | 0.06 | *84* | **0.05** | 0.03 | 0.06 | *26* | **0.04** | 0.03 | 0.05 | *30* | **0.04** | 0.03 | 0.06 | *17* | **0.045** | 0.04 | 0.065 | *12* |
| **Monocytes (giga/l)** | **0.47** | 0.37 | 0.59 | *84* | **0.43** | 0.35 | 0.64 | *26* | **0.465** | 0.3925 | 0.595 | *30* | **0.49** | 0.37 | 0.55 | *17* | **0.375** | 0.355 | 0.545 | *12* |
| **aPTT (sec)** | **29** | 27 | 31 | *85* | **29** | 27 | 31.925 | *26* | **29** | 28 | 30.75 | *30* | **29** | 27 | 32 | *18* | **30** | 29.025 | 31 | *12* |
| **INR** | **1** | 0.94 | 1 | *84* | **1** | 0.93 | 1 | *25* | **1** | 0.9925 | 1 | *30* | **1** | 1 | 1 | *18* | **1** | 1 | 1 | *12* |
| **D-Dimers (µg/ml)** | **0.28** | 0.19 | 0.49 | *83* | **0.29** | 0.21 | 0.49 | *25* | **0.235** | 0.19 | 0.3475 | *30* | **0.295** | 0.2 | 0.5 | *18* | **0.19** | 0.19 | 0.195 | *12* |
| **Antithrombin (%)** | **104** | 96 | 110.5 | *83* | **105** | 99 | 110 | *25* | **105.5** | 96 | 112.75 | *30* | **97** | 95 | 102.75 | *18* | **102** | 99.5 | 104.75 | *12* |
| **Fibrinogen (g/l)** | **2.75** | 2.41 | 3.26 | *83* | **2.84** | 2.68 | 3.22 | *25* | **2.54** | 2.2225 | 3.1375 | *30* | **2.77** | 2.535 | 3.345 | *18* | **2.285** | 2.1425 | 2.525 | *12* |
| **Factor VIII:c (%)** | **134.5** | 110.4 | 162 | *61* | **134.5** | 117.4 | 160.5 | *19* | **131** | 103 | 152.5 | *25* | **135.65** | 118.5 | 165.85 | *12* | **132.6** | 97.55 | 139.725 | *12* |
| **APC-Resistence** | **3.14** | 2.73 | 3.49 | *65* | **3.265** | 3.0675 | 3.795 | *20* | **3.16** | 2.97 | 3.49 | *25* | **2.99** | 2.655 | 3.305 | *15* | **3.295** | 3.24 | 3.505 | *10* |
| **Protein C (%)** | **108** | 94.6 | 121 | *83* | **111** | 99.25 | 122.75 | *26* | **111** | 94 | 125 | *29* | **95** | 90 | 111 | *17* | **96.5** | 87.5 | 103.5 | *10* |
| **Protein S (%)** | **96** | 81 | 110.5 | *65* | **96** | 87 | 109 | *19* | **96.5** | 82 | 114.25 | *26* | **87** | 80 | 99 | *13* | **89** | 80 | 96 | *12* |
| **LDH (U/l)** | **176** | 153 | 198 | *84* | **185** | 166 | 209 | *26* | **173** | 149 | 186 | *30* | **172.5** | 145 | 196.75 | *18* | **168** | 151.25 | 178.5 | *11* |
| **ALT (U/l)** | **20** | 15 | 28 | *86* | **19** | 17 | 25.5 | *27* | **20.5** | 15.5 | 34 | *30* | **18.5** | 14.25 | 24 | *18* | **18** | 15.5 | 21.5 | *12* |
| **AST (U/l)** | **21** | 18 | 26 | *86* | **23** | 18 | 26 | *27* | **21** | 17.25 | 23.5 | *30* | **21** | 17.5 | 24.5 | *18* | **16.5** | 14 | 21.25 | *12* |
| **Bilirubin (mg/dl)** | **0.6** | 0.5 | 0.8 | *85* | **0.6** | 0.5 | 0.9 | *27* | **0.5** | 0.4 | 0.7 | *30* | **0.695** | 0.525 | 0.775 | *18* | **0.8** | 0.7425 | 0.9 | *12* |
| **AP (U/l)** | **65** | 51 | 78.5 | *83* | **69** | 50.5 | 78.5 | *27* | **63.5** | 52.25 | 72 | *30* | **56.5** | 47.25 | 77.75 | *18* | **46** | 43 | 57.25 | *12* |
| **GGT (U/l)** | **16** | 12 | 22 | *82* | **15** | 12 | 21.5 | *27* | **17** | 12 | 26 | *29* | **16** | 11 | 19 | *18* | **16.5** | 12.5 | 19.25 | *12* |
| **HbA1c (%)** | **5.4** | 5.1 | 5.6 | *82* | **5.45315** | 5.3 | 5.5 | *27* | **5.30795** | 5 | 5.575 | *30* | **5.5** | 5 | 5.627 | *17* | **5.3** | 5 | 5.525 | *12* |
| **Creatinine (mg/dl)** | **0.7** | 0.7 | 0.8475 | *84* | **0.7** | 0.7 | 0.8 | *27* | **0.7** | 0.7 | 0.8775 | *30* | **0.8** | 0.7 | 0.9 | *18* | **0.77** | 0.7 | 0.85 | *12* |
| **Urea (mg/dl)** | **25** | 20 | 29.25 | *84* | **25.5** | 22 | 28.5 | *27* | **24.5** | 19.25 | 29.75 | *30* | **23** | 20 | 31 | *18* | **28.5** | 22 | 33.25 | *12* |
| **Uric acid (mg/dl)** | **4.8** | 4 | 5.6 | *83* | **4.6** | 3.9 | 5.3 | *25* | **5.2** | 4.325 | 6.05 | *30* | **4.45** | 4 | 5.4 | *18* | **4.7** | 4.25 | 5.75 | *12* |
| **Albumin (g/l)** | **45** | 43.575 | 47.325 | *81* | **44.5** | 42.5 | 45.6 | *25* | **46.05** | 43.8 | 47.8 | *30* | **44.7** | 44 | 47.5 | *17* | **44.5** | 43.625 | 46.35 | *11* |
| **Sodium (mmol/l)** | **139** | 138 | 139 | *86* | **139** | 138.5 | 140.5 | *27* | **139** | 138 | 140 | *30* | **139** | 138 | 140.75 | *18* | **140** | 138 | 140 | *12* |
| **Potassium (mmol/l)** | **4.1** | 3.8 | 4.3 | *85* | **3.9** | 3.7 | 4.1 | *27* | **4.1** | 3.9 | 4.275 | *30* | **4.15** | 3.925 | 4.3 | *18* | **3.95** | 3.875 | 4.025 | *12* |
| **Calcium (mmol/l)** | **2.3** | 2.2 | 2.4 | *83* | **2.2** | 2.2 | 2.35 | *27* | **2.3** | 2.2 | 2.3825 | *30* | **2.3** | 2.2 | 2.37 | *17* | **2.2** | 2.2 | 2.225 | *12* |
| **Magnesium (mmol/l)** | **0.83** | 0.79 | 0.88 | *83* | **0.8** | 0.7725 | 0.8775 | *26* | **0.815** | 0.7925 | 0.87 | *30* | **0.85** | 0.79 | 0.88 | *17* | **0.825** | 0.795 | 0.86 | *12* |
| **CRP (mg/l)** | **0.65** | 0.5 | 2.665 | *86* | **0.6** | 0.5 | 1.31 | *27* | **0.72** | 0.5 | 3.6275 | *30* | **0.5** | 0.5 | 2.1425 | *18* | **0.505** | 0.5 | 0.7125 | *12* |
| **CK (U/l)** | **90** | 56.75 | 138.5 | *84* | **108** | 75.25 | 154.25 | *26* | **78.5** | 54.5 | 132.25 | *30* | **87.5** | 52.75 | 104.75 | *18* | **140.5** | 92.5 | 243.25 | *12* |
| **BNP (pg/ml)** | **15** | 7 | 22 | *61* | **19.5** | 15.8 | 26.3 | *20* | **11** | 6.25 | 19.75 | *26* | **13** | 9 | 20 | *13* | **14.5** | 10 | 26.25 | *12* |
| **TNI (µg/l)** | **0.01** | 0.01 | 0.013 | *70* | **0.013** | 0.01 | 1.2905 | *23* | **0.01** | 0 | 0.0115 | *27* | **0.01** | 0 | 0.013 | *15* | **0.66** | 0.25 | 3.3 | *10* |
| **Ferritin (ng/ml)** | **49.75** | 24 | 92.25 | *80* | **51** | 37.2 | 112 | *25* | **48** | 24 | 94 | *29* | **41.25** | 17.75 | 78.5 | *18* | **41** | 12.75 | 85.175 | *9* |
| **Transferrin (g/l)** | **2.365** | 2.18 | 2.6 | *72* | **2.27** | 2.165 | 2.415 | *23* | **2.43** | 2.1525 | 2.6175 | *28* | **2.46** | 2.275 | 2.7 | *16* | **2.38** | 2.155 | 2.6425 | *12* |
| **ACE (U/l)** | **36** | 25.7 | 55.5 | *79* | **36.8** | 24 | 59 | *25* | **35** | 26 | 51 | *29* | **40.5** | 31.75 | 64.25 | *18* | **29** | 24.5 | 34.5 | *10* |
| **sIL-2 (U/ml)** | **332** | 272 | 434 | *70* | **394** | 266 | 476 | *21* | **318** | 287.75 | 385.25 | *28* | **310** | 265.5 | 385 | *15* | **220.5** | 193 | 265 | *11* |
| **Cholesterol (mg/dl)** | **190** | 159 | 223 | *81* | **201** | 176 | 233 | *26* | **194** | 163.5 | 222.25 | *30* | **177** | 147 | 209 | *17* | **166.5** | 146.25 | 190 | *10* |
| **HDL (mg/dl)** | **56** | 43.5 | 65 | *83* | **61** | 53 | 70.5 | *27* | **51.5** | 41 | 59 | *30* | **55** | 41 | 64 | *17* | **56** | 48.75 | 66 | *12* |
| **LDL (md/dl)** | **129** | 95.5 | 166.75 | *82* | **146** | 106.5 | 167 | *27* | **124** | 100.5 | 169.75 | *30* | **105** | 84 | 150 | *17* | **104.5** | 90 | 122.5 | *12* |
| **Triglyceride (mg/dl)** | **93.5** | 64.25 | 149 | *82* | **76** | 61 | 119 | *27* | **132** | 71.25 | 176 | *30* | **76** | 58 | 134 | *17* | **50** | 44.5 | 67 | *12* |
| **Lipase (U/l)** | **35** | 30.5 | 43 | *83* | **35** | 30.75 | 44.5 | *26* | **35.5** | 33.25 | 41 | *30* | **35** | 29 | 45 | *18* | **36.5** | 32 | 49 | *11* |
| **Amylase (U/l)** | **57** | 46 | 76 | *81* | **56** | 50 | 76.75 | *26* | **56.5** | 41.25 | 65.75 | *30* | **64** | 54 | 76 | *17* | **60** | 47.5 | 65.5 | *10* |
| **IgE (U/ml)** | **24.3** | 7.45 | 66.5 | *79* | **25.5** | 6.2 | 69.1 | *26* | **24.3** | 5.8 | 42.5 | *29* | **17.1** | 15 | 50.1 | *17* | **28** | 11.4 | 71.6 | *11* |
| **IgG (g/l)** | **10.35** | 8.775 | 11.8 | *72* | **9.9** | 8 | 11.8 | *21* | **10.3** | 8.9 | 11.7 | *29* | **10.4** | 9.475 | 10.975 | *16* | **9** | 8.65 | 10.225 | *9* |
| **25-OH-Vitamin D3 (ng/ml)** | **24.4** | 17.3 | 33.1 | *77* | **25.5** | 18.5 | 42.3 | *25* | **21.9** | 15.2 | 29.9 | *29* | **26.8** | 20.5 | 36.7 | *15* | **26.1** | 10.6 | 28.1 | *10* |
| **TSH (mU/l)** | **1.91** | 1.255 | 2.73 | *83* | **1.585** | 1.125 | 2.545 | *26* | **2.4** | 1.44 | 3.14 | *29* | **2.235** | 1.3825 | 2.35 | *18* | **1.42** | 1.2 | 2.12 | *9* |
| **fT4 (ng/dl)** | **1.2** | 1.1 | 1.3 | *70* | **1.2** | 1.1 | 1.3 | *23* | **1.1** | 1 | 1.2 | *27* | **1.2** | 1.1 | 1.3 | *16* | **1.3** | 1 | 1.425 | *11* |
| **CD4+ T cells (%)** | **49** | 45 | 53 | *60* | **50** | 48.5 | 53 | *20* | **47** | 45 | 50.75 | *26* | **49** | 43 | 55 | *13* | | | | *0* |
| **CD8+ T cells (%)** | **24.5** | 20.5 | 29 | *60* | **24** | 18.5 | 28 | *19* | **26** | 21.25 | 28.75 | *26* | **22** | 21 | 26 | *13* | | | | *8* |
| **Folic acid (ng/ml)** | **13.85** | 9.475 | 18.725 | *56* | **14.7** | 10.1 | 19.9 | *17* | **13.3** | 8.475 | 15.575 | *26* | **14.6** | 9.7 | 34.65 | *10* | **13.4** | 10.375 | 19.55 | *0* |
| **Vit B12 (pg/ml)** | **459** | 354 | 548 | *57* | **396** | 354 | 590 | *17* | **447** | 328 | 543 | *26* | **472** | 399 | 536 | *11* | **425** | 389 | 581 | *8* |

*Supplemental Table S9*: Lab values and findings parameters at rest for groups: All patients, patients with normal peak$VO_2$ (Group 1), patients with reduced peak$VO_2$ and maximal effort (Group 2), patients with reduced peak$VO_2$ (Group 3) and control group.

*Hb: hemoglobin, Hct: hematocrit, RBC: red blood cells, Platelets: platelets, WBC: white blood cells, MCV: mean corpuscular volume, MCH: mean corpuscular hemoglobin, Neutrophils: neutrophils, Lymphocytes: lymphocytes, Eosinophils: eosinophils, Basophils: basophils, Monocytes: monocytes, aPTT: activated partial thromboplastin time, INR : international normalized ratio, D-Dimers: d-dimers, Antithrombin: antithrombin, Fibrinogen: fibrinogen, VIII:c: factor VIII, APC-Resistance: activated protein C resistance, C Protein: protein C, Protein S: protein S, LDH: lactate dehydrogenase, ALT: alanine aminotransferase, AST: aspartate aminotransferase, Bilirubin: bilirubin, AP: alkaline phosphatase, GGT: gamma-glutamyl transferase, HbA1c: hemoglobin a1c, Creatinine: creatinine, Urea: urea, Uric acid: uric acid, Albumin: albumin, Sodium: sodium, Potassium: potassium, Calcium: calcium, Magnesium: magnesium, CRP: c-reactive protein, CK: creatine kinase, BNP: b-type natriuretic peptide, TNI: troponin I, Ferritin: ferritin, Transferrin: transferrin, ACE: angiotensin-converting enzyme, sIL-2: soluble interleukin-2, Cholesterol: cholesterol, HDL: high-density lipoprotein, LDL: low-density lipoprotein, Triglyceride: triglycerides, Lipase: lipase, Amylase: amylase, IgE: immunoglobulin E, IgG: immunoglobulin G, 25-OH-vitamin D3: 25-hydroxyvitamin D3, TSH: thyroid-stimulating hormone, fT4: free thyroxine, CD4+ T cells: CD4+ T cells, CD8+ T cells: CD8+ T cells, Folic acid: folic acid, Vit B12: vitamin B12*

| | All patients | Normal peak$VO_2$ (Group 1) | Low peak$VO_2$, maximal effort (Group 2) | Low peak$VO_2$, submaximal effort (Group 3) | Control |
|---|---|---|---|---|---|
| Antikoagulation | 4 | 2 | 1 | 1 | 0 |
| ACE inhibitors | 7 | 1 | 4 | 1 | 1 |
| Beta blocker | 16 | 4 | 6 | 5 | 1 |
| Diuretics | 2 | 1 | 1 | 0 | 0 |
| Inhalatives | 14 | 6 | 3 | 5 | 0 |
| Immunosuppressants | 3 | 1 | 2 | 0 | 0 |
| Antidepressants | 8 | 3 | 3 | 2 | 0 |
| Other Medication* | 21 | 6 | 8 | 5 | 2 |

*e.g. Thyroid hormones, Biguanides, Statins (HMG-CoA reductase inhibitors), Analgesics, Heparins, Vitamins

*Supplemental Table S10:* Overview of the medications taken by the patients in the respective groups
