## Supplemental Fig. for "Preload insufficiency as common denominator of exertional dyspnoea in distinct post-COVID phenotypes"

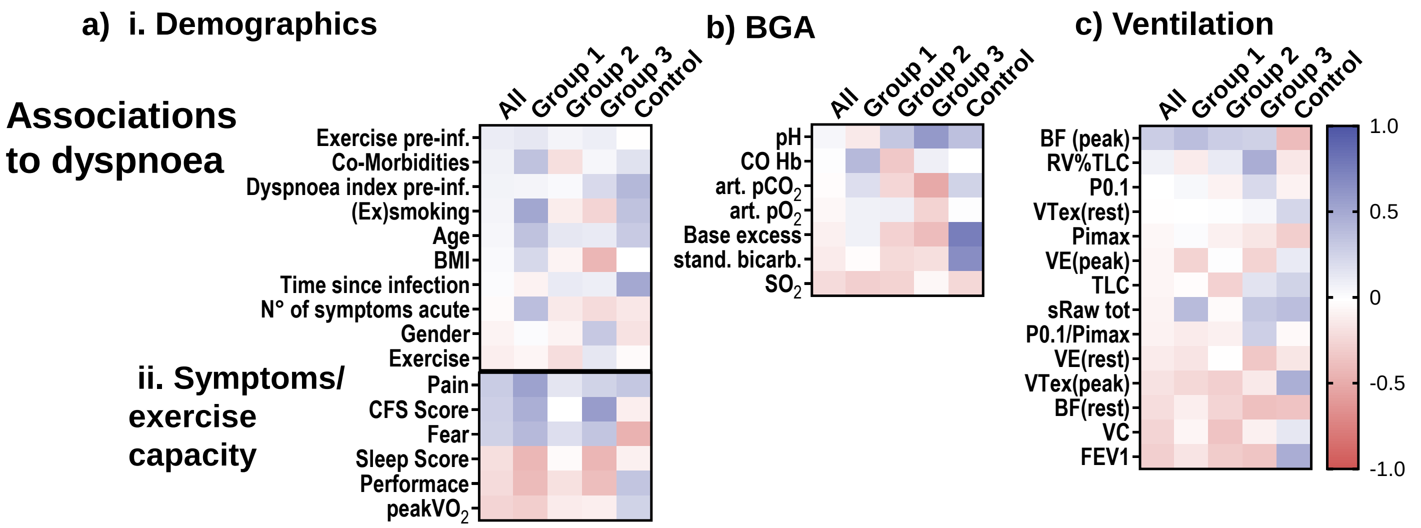


| …in all patients | Other symptoms low exercise capacity | Low oxygenation High heart rate | Low lung volumes high breathing frequence |
|---|---|---|---|
| ..in patients with cardiac or ventilatory limitation and normal peak$VO_2$ (G1) | (Ex-)smoking | High CO-Hb (as e.g. in smokers) | Increased airway resistance |
| in patients with cardiac or ventilatory limitation and low peak$VO_2$ (G2) | | | |
| in patients without cardiac or ventilatory imitation (G3) | Low BMI | Chronic hyperventilation | High RV%TLC |

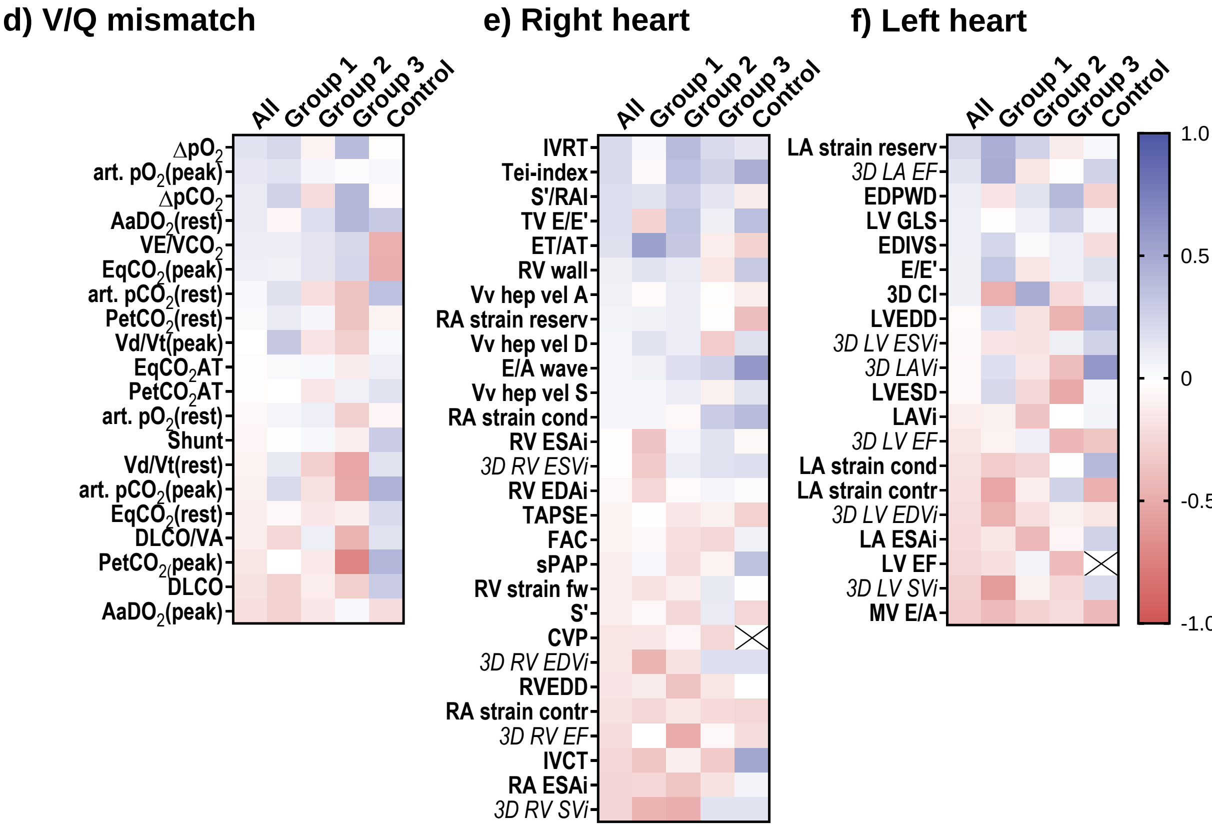


| …in all patients | DLCO (% pred) | Low volumes, low function | Diastolic dysfunction |
|---|---|---|---|
| ..in patients with cardiac or ventilatory limitation and normal peak$VO_2$ (G1) | Signs of V/Q mismatch | | Low function |
| in patients with cardiac or ventilatory limitation and low peak$VO_2$ (G2) | | High CO-PA | High CI |
| in patients without cardiac or ventilatory imitation (G3) | Signs of V/Q mismatch Hyperventilation | | |

Supplementary Fig. 1: Association of dyspnoea to a) demographics/symptoms, b) vital parameters, c) ventilation, d) V/Q mismatch, e) right heart parameters and f) left heart parameters for all patients, different patient groups and controls (Co). Heatmaps represent pearsons correlation coefficent sorted for all patients.

Continued on the following page.

*Suppl. Figure 1:*

*f/m: female/male, BMI: body mass index, ME/CFS Score: myalgic encephalomyelitis/chronis fatigue score, CFS score: clinical frailty scale score, N°: number. pre-inf.: pre-infection, peak$VO_2$: oxygen uptake at peak exercise. pred.: predicted, $SO_2$: oxygen saturation, art.: arterial, art. $pCO_2$: partial pressure of carbon dioxide, art. $pO_2$: partial pressure of oxygen. VC: vital capacity. FEV1: forced expiratory volume in 1 s, sRaw tot: total specific resistance, RV%TLC: residual volume in % total lung capacity, TLC: total lung capacity, CO Hb: Carboxyhaemoglobin, P0.1: airway occlusion pressure, Pimax: maximal inspiratory pressure, VE: ventilation, BF: Breath rate, ΔpO2: venous-to-arterial oxygen difference, ΔpCO2: venous-to-arterial carbon dioxide, DLCO: diffusing capacity for carbon monoxide, DLCO/VA: diffusing capacity/Alveolar volume, EqCO2: ventilatory equivalent of carbon dioxide, VE/VCO2: ventilation/carbon dioxide production, PetCO2: end-tidal carbon dioxide tension, VDc/VT: ratio of physiologic dead space over tidal volume, AaDO2: alveolar-arterial oxygen gradient, sPAP: systolic pulmonary artery pressure. TAPSE: tricuspid annular plane systolic excursion. Tei-index: myocardial performance index, ET/AT: ejection time/acceleration time, FAC: fractional area change, IVCT: isovolumic contraction time, IVRT: isovolumic relaxation time, S‘: systolic contraction velocity, S‘/RAi: systolic contraction velocity/right atrial index, TV: tricuspid valve, RVEDD: right ventricular end-diastolic diameter, E/A wave: early to late diastolic transtricuspid flow velocity, E/E‘: ratio of the E wave velocity to myocardial movement velocity. RA: right atrium, ESAi: end-systolic area index, RV: right ventricular, CVP: central venous pressure, EDD: end-diastolic diameter, EDAi: end-diastolic area index, ESAi: end-systolic area index, EDVi: end-diastolic volume index, ESVi: end-systolic volume index, SVi: stroke volume index, EF: ejection fraction, LV EF: left ventricular ejection fraction, MV E/A: mitral valve E-wave/A-wave ratio, E/E‘: left ventricular filling pressure, EDIVS: end.diastolic volume of the left heart, EDPWD: end-diastolic posterior wall diameter, LA ESAi: left atrial end systolic area, LAVI: left atrial volume index, EDPWD: , EDIVS: , LVEDD: left ventricular end-diastolic diameter, LVESD: left ventricular end-systolic diameter*, *LA strain Reserv: left atrial strain reservoir, LA strain Cond: left atrial strain conduit, LA strain Kontr: left atrial strain conduit, 3D LV EDVi: 3D Left ventricular end-diastolic volume, 3D LV ESVI: 3D Left ventricular end-systolic volume, 3D LV SVi: 3D Left ventricular stroke volume, 3D CI: 3D Cardiac index, 3D LV EF: 3D Left ventricular ejection fraction, LV GLS Strain: left ventricular global longitudinal strain*

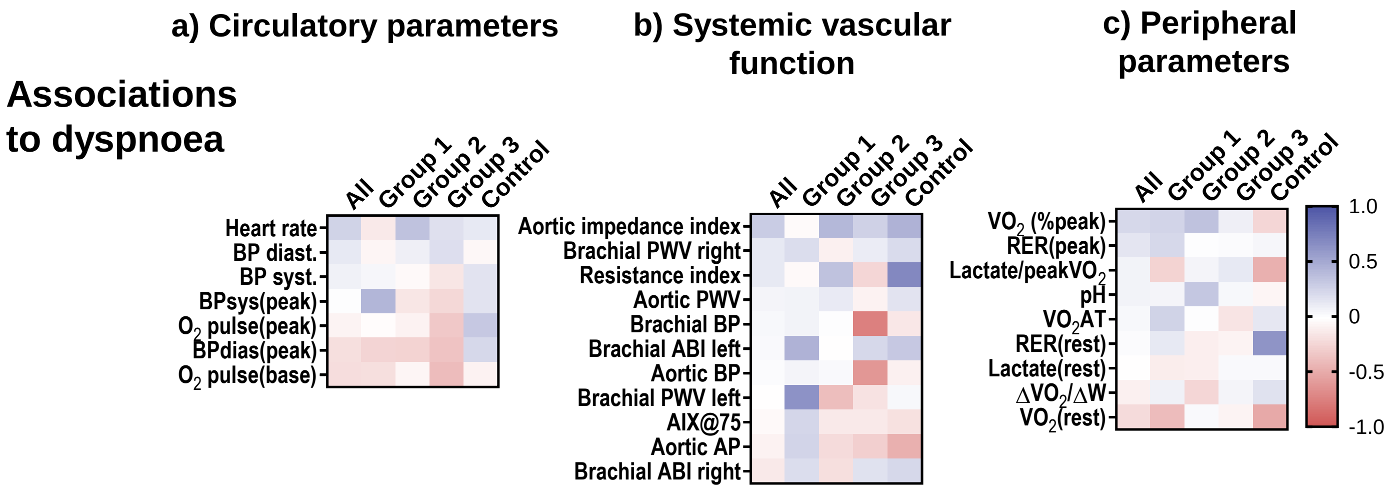


| ...in all patients | | | Low oxygen extraction |
|---|---|---|---|
| ..in patients with cardiac or ventilatory limitation and normal peak$VO_2$ (G1) | High systolic blood pressure | High blood pressure and pulse wave velocity | |
| in patients with cardiac or ventilatory limitation and low peak$VO_2$ (G2) | High heart rate | High heart rate | |
| in patients without cardiac or ventilatory imitation (G3) | Low blood pressure | Low blood pressure | |

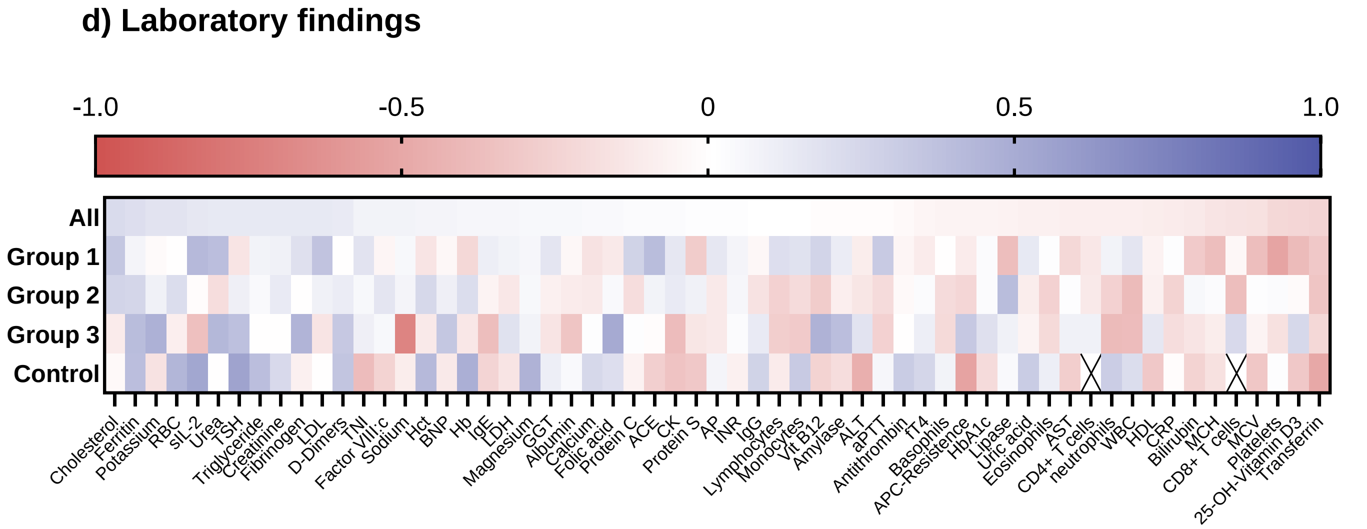


Supplementary Fig. 2: Association of dyspnea to a) circulatory parameters, b) systemic vascular parameters, c) peripheral parameters, d) laboratory findings for all patients, different patient groups and controls (Co). Heatmaps represent pearsons correlation coefficent sorted for all patients.

*HF: heart rate, BP: blood prssure, sys: systolic, dias: diastolic, ΔVO2/ΔW: peak oxygen consumption to watt relationship, PWV: pulse wave velocity, BP: blood pressure. AIX(%): augmentation index, AIX@75: augumentation index normalized to a heart rate of 75 beats per minute, ABI: Ancle-brachial-index,* RER: respiratory exchange ratio VO2AT: Oxygen consumption at anaerobic threshold, ΔVO2/ΔW: peak oxygen consumption to watt relationship, *Hb: hemoglobin, Hct: hematocrit, RBC: red blood cells, Platelets: platelets, WBC: white blood cells, MCV: mean corpuscular volume, MCH: mean corpuscular hemoglobin, Neutrophils: neutrophils, Lymphocytes: lymphocytes, Eosinophils: eosinophils, Basophils: basophils, Monocytes: monocytes, aPTT: activated partial thromboplastin time, INR : international normalized ratio, D-Dimers: d-dimers, Antithrombin: antithrombin, Fibrinogen: fibrinogen, VIII:c: factor VIII, APC-Resistance: activated protein C resistance, C Protein: protein C, Protein S: protein S, LDH: lactate dehydrogenase, ALT: alanine aminotransferase, AST: aspartate aminotransferase, Bilirubin: bilirubin, AP: alkaline phosphatase, GGT: gamma-glutamyl transferase, HbA1c: hemoglobin a1c, Creatinine: creatinine, Urea: urea, Uric acid: uric acid, Albumin: albumin, Sodium: sodium, Potassium: potassium, Calcium: calcium, Magnesium: magnesium, CRP: c-reactive protein, CK: creatine kinase, BNP: b-type natriuretic peptide, TNI: troponin I, Ferritin: ferritin, Transferrin: transferrin, ACE: angiotensin-converting enzyme, sIL-2: soluble interleukin-2, Cholesterol: cholesterol, HDL: high-density lipoprotein, LDL: low-density lipoprotein, Triglyceride: triglycerides, Lipase: lipase, Amylase: amylase, IgE: immunoglobulin E, IgG: immunoglobulin G, 25-OH-vitamin D3: 25-hydroxyvitamin D3, TSH: thyroid-stimulating hormone, fT4: free thyroxine, CD4+ T cells: CD4+ T cells, CD8+ T cells: CD8+ T cells, Folic acid: folic acid, Vit B12: vitamin B12*

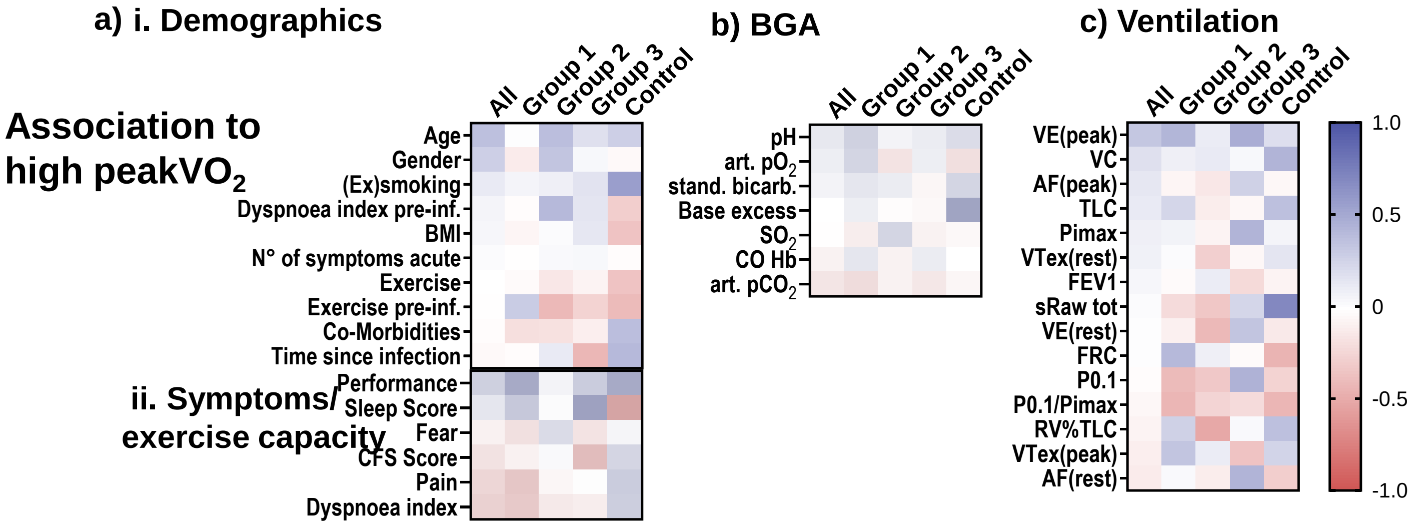


| …in all patients | High age, low dyspnea | Low heart rate | |
|---|---|---|---|
| ..in patients with cardiac or ventilatory limitation and normal peak$VO_2$ (G1) | | High oxygenation, low pCO2 | High lung volumes |
| in patients with cardiac or ventilatory limitation and low peak$VO_2$ (G2) | low exercise and high dyspnea before infection | | Low hyperinflation |
| in patients without cardiac or ventilatory imitation (G3) | | | High inspiratory strength |

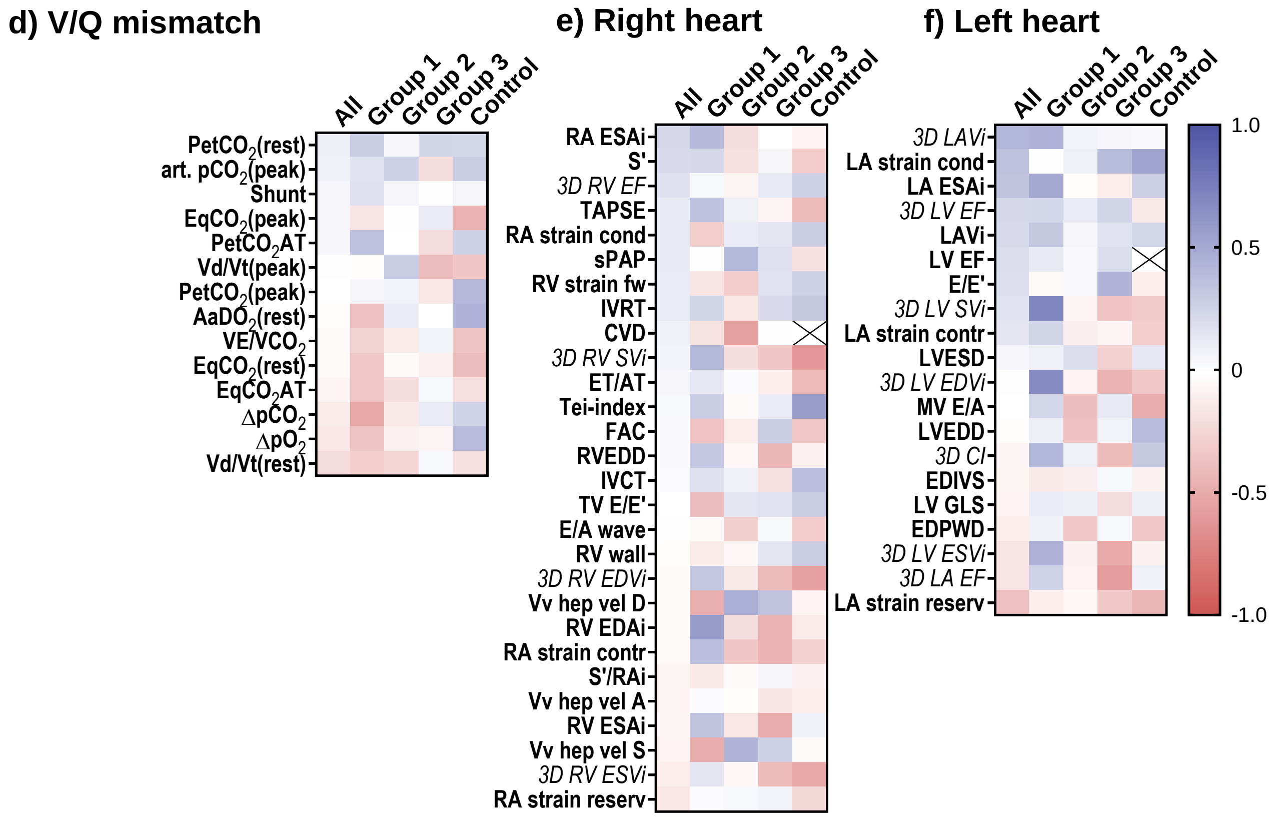


| …in all patients | | High volumes and function | |
|---|---|---|---|
| ..in patients with cardiac or ventilatory limitation and normal peak$VO_2$ (G1) | High diffusion and $pO_2$, low V/Q mismatch | High RA area, stroke volume | High function and volumes |
| in patients with cardiac or ventilatory limitation and low peak$VO_2$ (G2) | | Low CVP | |
| in patients without cardiac or ventilatory imitation (G3) | Low dead space | | Low function |

Supplementary Fig. 3: Association of peak $VO_2$ to a) demographics, b) vital parameters, c) ventilation, d) V/Q mismatch, e) right heart parameters and f) left heart parameters for all patients, different patient groups and controls (Co). Heatmaps represent pearsons correlation coefficent sorted for all patients.

Continued on the following page.

*Suppl. Figure 3:*

*f/m: female/male, BMI: body mass index, ME/CFS Score: myalgic encephalomyelitis/chronis fatigue score, CFS score: clinical frailty scale score, N°: number. pre-inf.: pre-infection, peak$VO_2$: oxygen uptake at peak exercise. pred.: predicted, $SO_2$: oxygen saturation, art.: arterial, art. $pCO_2$: partial pressure of carbon dioxide, art. $pO_2$: partial pressure of oxygen. VC: vital capacity. FEV1: forced expiratory volume in 1 s, sRaw tot: total specific resistance, RV%TLC: residual volume in % total lung capacity, TLC: total lung capacity, CO Hb: Carboxyhaemoglobin, P0.1: airway occlusion pressure, Pimax: maximal inspiratory pressure, VE: ventilation, BF: Breath rate, ΔpO2: venous-to-arterial oxygen difference, ΔpCO2: venous-to-arterial carbon dioxide, DLCO: diffusing capacity for carbon monoxide, DLCO/VA: diffusing capacity/Alveolar volume, EqCO2: ventilatory equivalent of carbon dioxide, VE/VCO2: ventilation/carbon dioxide production, PetCO2: end-tidal carbon dioxide tension, VDc/VT: ratio of physiologic dead space over tidal volume, AaDO2: alveolar-arterial oxygen gradient, sPAP: systolic pulmonary artery pressure. TAPSE: tricuspid annular plane systolic excursion. Tei-index: myocardial performance index, ET/AT: ejection time/acceleration time, FAC: fractional area change, IVCT: isovolumic contraction time, IVRT: isovolumic relaxation time, S‘: systolic contraction velocity, S‘/RAi: systolic contraction velocity/right atrial index, TV: tricuspid valve, RVEDD: right ventricular end-diastolic diameter, E/A wave: early to late diastolic transtricuspid flow velocity, E/E‘: ratio of the E wave velocity to myocardial movement velocity. RA: right atrium, ESAi: end-systolic area index, RV: right ventricular, CVP: central venous pressure, EDD: end-diastolic diameter, EDAi: end-diastolic area index, ESAi: end-systolic area index, EDVi: end-diastolic volume index, ESVi: end-systolic volume index, SVi: stroke volume index, EF: ejection fraction, LV EF: left ventricular ejection fraction, MV E/A: mitral valve E-wave/A-wave ratio, E/E‘: left ventricular filling pressure, EDIVS: end.diastolic volume of the left heart, EDPWD: end-diastolic posterior wall diameter, LA ESAi: left atrial end systolic area, LAVI: left atrial volume index, EDPWD: , EDIVS: , LVEDD: left ventricular end-diastolic diameter, LVESD: left ventricular end-systolic diameter*, *LA strain Reserv: left atrial strain reservoir, LA strain Cond: left atrial strain conduit, LA strain Kontr: left atrial strain conduit, 3D LV EDVi: 3D Left ventricular end-diastolic volume, 3D LV ESVI: 3D Left ventricular end-systolic volume, 3D LV SVi: 3D Left ventricular stroke volume, 3D CI: 3D Cardiac index, 3D LV EF: 3D Left ventricular ejection fraction, LV GLS Strain: left ventricular global longitudinal strain*

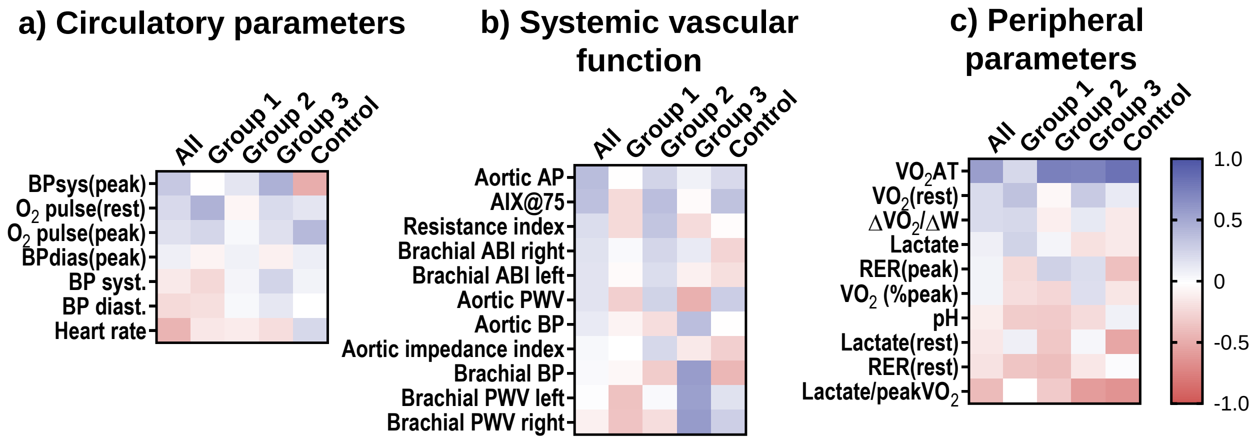


| …in all patients | | | High aerobic treshold, low lactate |
|---|---|---|---|
| ..in patients with cardiac or ventilatory limitation and normal peak$VO_2$ (G1) | High oxygen pulse | Low pulse wave velocities | |
| in patients with cardiac or ventilatory limitation and low peak$VO_2$ (G2) | | High calendar age | |
| in patients without cardiac or ventilatory imitation (G3) | High blood pressure | High pulse wave velocities | |

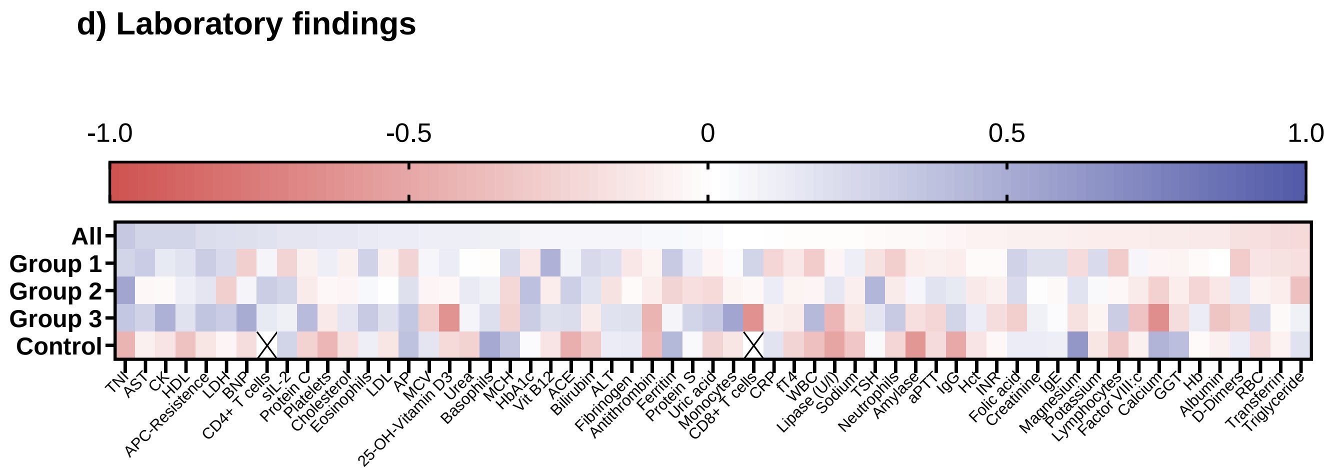


Supplementary Fig. 4: Association of peak $VO_2$ to a) circulatory parameters, b) systemic vascular parameters, c) peripheral parameters, d) laboratory findings for all patients, different patient groups and controls (Co). Heatmaps represent pearsons correlation coefficent sorted for all patients.

*HF: heart rate, BP: blood prssure, sys: systolic, dias: diastolic, ΔVO2/ΔW: peak oxygen consumption to watt relationship, PWV: pulse wave velocity, BP: blood pressure. AIX(%): augmentation index, AIX@75: augumentation index normalized to a heart rate of 75 beats per minute, ABI: Ancle-brachial-index*, RER: respiratory exchange ratio VO2AT: Oxygen consumption at anaerobic threshold, ΔVO2/ΔW: peak oxygen consumption to watt relationship, *Hb: hemoglobin, Hct: hematocrit, RBC: red blood cells, Platelets: platelets, WBC: white blood cells, MCV: mean corpuscular volume, MCH: mean corpuscular hemoglobin, Neutrophils: neutrophils, Lymphocytes: lymphocytes, Eosinophils: eosinophils, Basophils: basophils, Monocytes: monocytes, aPTT: activated partial thromboplastin time, INR : international normalized ratio, D-Dimers: d-dimers, Antithrombin: antithrombin, Fibrinogen: fibrinogen, VIII:c: factor VIII, APC-Resistance: activated protein C resistance, C Protein: protein C, Protein S: protein S, LDH: lactate dehydrogenase, ALT: alanine aminotransferase, AST: aspartate aminotransferase, Bilirubin: bilirubin, AP: alkaline phosphatase, GGT: gamma-glutamyl transferase, HbA1c: hemoglobin a1c, Creatinine: creatinine, Urea: urea, Uric acid: uric acid, Albumin: albumin, Sodium: sodium, Potassium: potassium, Calcium: calcium, Magnesium: magnesium, CRP: c-reactive protein, CK: creatine kinase, BNP: b-type natriuretic peptide, TNI: troponin I, Ferritin: ferritin, Transferrin: transferrin, ACE: angiotensin-converting enzyme, sIL-2: soluble interleukin-2, Cholesterol: cholesterol, HDL: high-density lipoprotein, LDL: low-density lipoprotein, Triglyceride: triglycerides, Lipase: lipase, Amylase: amylase, IgE: immunoglobulin E, IgG: immunoglobulin G, 25-OH-vitamin D3: 25-hydroxyvitamin D3, TSH: thyroid-stimulating hormone, fT4: free thyroxine, CD4+ T cells: CD4+ T cells, CD8+ T cells: CD8+ T cells, Folic acid: folic acid, Vit B12: vitamin B12*

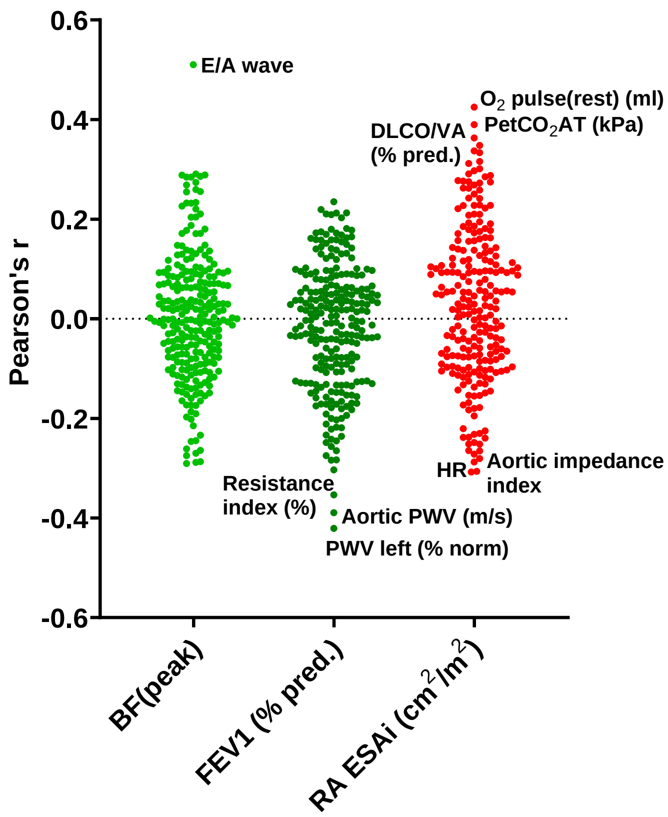


Supplmentary Fig. 5: Associations of breathing frequency at peak exercise (BF peak), forced expiratory volume at 1 s (FEV1) and right atrial end systolic area index (RA ESAi) with other cardiopulmonary parameters

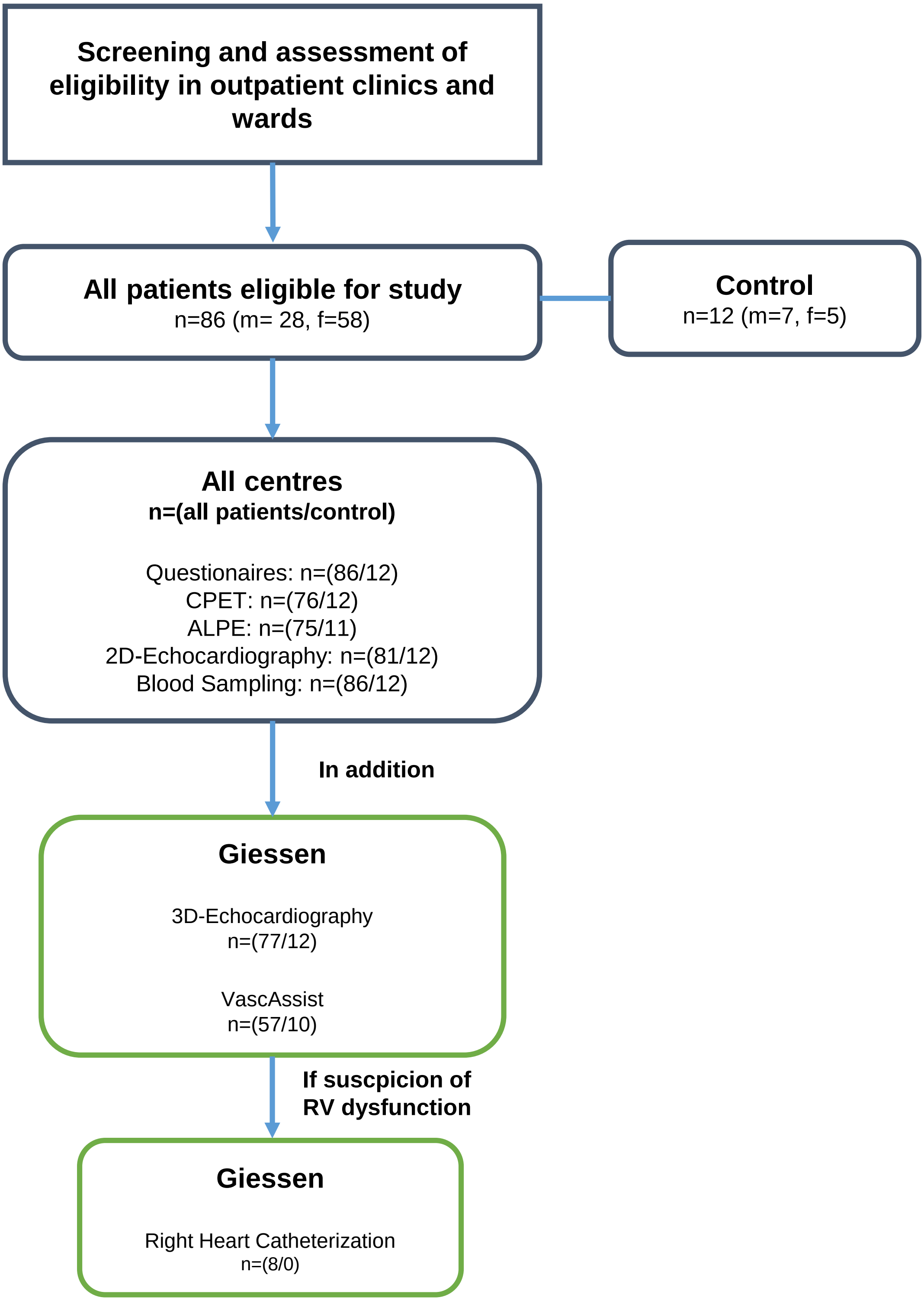


Supplementary Fig. 6: Methodical flow chart including study population and methods of the multicenter, cross sectional study, conducted between 02/2022 and 02/2024.
