## Supplemental Methods for "Preload insufficiency as common denominator of exertional dyspnoea in distinct post-COVID phenotypes"

Study Design and Study Population

PulmVasC was designed as multicenter, cross-sectional study conducted between 02/2022 and 02/2024 at four tertiary care centers in Germany, including Justus-Liebig University of Giessen, Charité – Universitätsmedizin Berlin, Hanover Medical School, and Ludwig-Maximilian University Munich. A total of 98 participants aged ≥18 years were enrolled, comprising 86 patients and 12 healthy controls. Patients included in the study had a confirmed history of SARS-CoV-2 infection and were referred to a specialized post-COVID-19 outpatient clinics for unexplained dyspnoea persisting or developing after more than three months after the acute infection according to the WHO definition of the post-COVID condition. Controls were individuals with a documented SARS-CoV-2 infection and no chronic symptoms. All participants underwent a comprehensive outpatient assessment that included standardized symptom questionnaires, pulmonary function testing, blood gas analysis, resting and exercise echocardiography, and cardiopulmonary exercise testing (CPET) [1]. In a subset of patients, right heart catheterization was performed based on clinical indication. In addition, venous blood was collected from each participant for further analysis. Written informed consent was obtained from all participants prior to study inclusion. The study was approved by the local ethics committees at all participating institutions (lead ethics committee: Faculty of Medicine, Justus Liebig University Giessen, reference number AZ 28/22).

Blood Sampling and Testing

Fasting venous blood samples were collected from each participant for subsequent analysis. Routine clinical laboratory parameters were measured using standardized protocols at the Institute of Laboratory Chemistry and Pathobiology, University Hospital Giessen (Germany).

Questionnaires

At study inclusion, all participants completed a series of standardized and study-specific questionnaires. These were designed to assess general health status, symptom burden, and functional impairment, with a focus on fatigue, cognitive function, mental health, and social reintegration. Fatigue, a common and debilitating symptom in post-COVID-19 recovery, was evaluated using the Chandler Fatigue Scale (CFS). A short overview of medication taken by the patients are listed in supplemental data (Supplemental Table 11). Additional self-designed instruments captured patient-reported outcomes specific to post-COVID-19 symptomatology.

Pulmonary Function Test

All participants underwent standardized pulmonary function testing (PFT) in accordance with current international guidelines [2], including measurement of static and dynamic lung volumes as well as diffusing capacity of the lung for carbon monoxide (DLCO). PFTs were performed using body plethysmography (Vyntus™ BODY, CareFusion, Höchberg, Germany). Parameters were recorded and expressed as percentages of predicted normal values, adjusted for age, sex, height, and weight. DLCO values were corrected for hemoglobin concentration when available.

Ventilation-Perfusion Analysis: Automatic Lung Parameter Estimator (ALPE)

Ventilation-perfusion mismatch was assessed using the Automatic Lung Parameter Estimator (ALPE2, Mermaid Care A/S, Stenlose, Denmark), a non-invasive system designed for spontaneously breathing patients to quantify key parameters of pulmonary gas exchange [3]. The ALPE method estimates gas exchange abnormalities, including pulmonary shunt and ventilation-perfusion (V/Q) mismatch, through a series of controlled step changes in the fraction of inspired oxygen (FiO₂) and a mathematical two-compartment model of lung physiology. Each participant underwent ALPE testing while breathing through a mouthpiece connected to the respiratory unit, with their nose securely sealed to ensure exclusive oral breathing [4]. A bacterial and humidity filter was used to ensure measurement integrity. During the procedure, oxygen was delivered continuously and adjusted in 2-3 incremental steps, typically ranging from inspired oxygen concentrations of 21% to 30% (0-3 L/min O₂). Each step was maintained until a steady state was achieved, generally within 5 minutes. Endtidal gas values were measured breath-by-breath, and capillary blood gases obtained. The system computed pulmonary shunt fraction as well as indices of low and high V/Q mismatch, namely ΔpO₂ and ΔpCO₂, respectively. ΔpO₂ and ΔpCO₂ reflect the difference of measured blood gases and calculated blood gases for ideal V/Q matching.

Cardiopulmonary Exercise Testing (CPET)

Cardiopulmonary exercise testing was performed in a semi-recumbent position using an electronically braked bicycle ergometer (Ergoline, Ergoline GmbH, Bitz, Germany; Vyntus PNEUMO, CareFusion, Höchberg, Germany) to assess peak oxygen uptake (peakVO₂) in accordance with current international guidelines [1]. Each test followed an individualized ramp protocol, with the ramp rate tailored to the participant’s physical fitness or activity level. After a 3-minute warm-up phase at 0 watts (W), the workload was incrementally increased by 10 W every 3 minutes until the participant reached their individual limit of exercise tolerance. All participants underwent breath-by-breath gas exchange analysis throughout the test, including continuous monitoring of ventilation (V̇E), tidal volume, respiratory rate, oxygen saturation (SpO₂), end-tidal oxygen (PetO₂) and carbon dioxide (PetCO₂) concentrations. Different parameters were calculated such as oxygen uptake (V̇O₂), carbon dioxide production (V̇CO₂), oxygen pulse, and ventilatory equivalents for oxygen (V̇E/V̇O₂) and carbon dioxide (V̇E/V̇CO₂). Cardiac function was concurrently assessed at rest and during peak exercise using transthoracic echocardiography.

Echocardiography (2D and 3D)

Comprehensive transthoracic echocardiography, including Doppler, speckle-tracking strain, and three-dimensional (3D) imaging, was performed in accordance with current international guidelines [5] and as previously described [6]. Examinations were conducted using a Philips EPIQ 7G ultrasound system (Philips Healthcare, the Netherlands), with participants in the left lateral position. Three-dimensional volumetric imaging was acquired in dedicated right and left ventricular apical views, ensuring full visualization of the endocardial borders. End-diastolic volume (EDV) and end-systolic volume (ESV) were measured directly, from which stroke volume (SV) was calculated as EDV – ESV, and ejection fraction (EF) as (EDV – ESV)/EDV. Volumes were indexed to body surface area (BSA) where appropriate. 3D left atrial volumetry was obtained by the implemented DHM software (Dynamic HeartModelA.I., Philips Healthcare). Strain measurements and 2-dimensional functional and dimensional parameters were obtained as previously described [7].

Noninvasive Assessment of Peripheral, Central Blood Pressure and Pulse Pressure Waveforms

The Vascassist2® device (iSYMED GmbH, Butzbach, Germany) was used for the noninvasive measurement of pulse pressure waveforms through oscillometry as previous reported [8]. In short, the pulse pressure waveforms were analyzed to calculate key vascular parameters, including brachial and radial blood pressures, central blood pressures, aortic pulse wave velocity (PWV), augmentation index (Aix), Aix at 75 bpm, resistance index, and ejection duration. All participants underwent the noninvasive vascular evaluation after a 15-minute rest period. Measurements were taken in a supine position using four standard cuffs, adjusted to fit the participants’ arm and forearm circumferences. Pulse pressure waves from both radial and brachial arteries were recorded on each arm through gradual deflation of the cuffs. Central blood pressure was calculated using a transfer function based on the peripheral waveform, while Aix at a heart rate of 75 was derived from the pulse waveform.

Right Heart Catheterization (RHC)

RHC was performed due to clinical indication (e.g. signs of right heart dysfunction in echocardiography). Hemodynamic parameters were measured by a Swan-Ganz catheter: mean pulmonary artery pressure (mPAP), central venous pressure (CVP), right ventricular (RV) pressure, and pulmonary artery wedge pressure (PAWP). Cardiac output (CO) was determined using either the using direct Fick (during rest) or thermodilution (during rest and exercise) in all patients. Cardiac index (CI) was calculated as the ratio of CO to the calculated body surface area (BSA). BSA was determined using the formula provided by DuBois: BSA = 0.007184 × height^0.725 × weight^0.425. Pulmonary vascular resistance (PVR) was calculated as: PVR = (mPAP – PAWP) / CO [7].

**Statistical analysis**

Data collection was conducted using RedCap electronic data capture tools, and all statistical analyses were performed using GraphPad Prism (version 9.0.2). For comparisons between patients and control participants Student’s t-test was used for normally distributed values, while Mann-Whitney was applied for non-normally distributed data. Simple linear regression analyses were performed for all parameters, and multivariate linear regression for potential confounders (age, gender) and parameters with a r>0.25 or r<-0.25 in the linear regression. To evaluate associations between different functional clusters to dyspnoea or peakVO_2_, Fisher’s r-to-z transformation was applied for each r-value of a parameter in a cluster, the transformed absolute z-values from one cluster averaged and re-transformed to r-values.

**References**

1.	Balady GJ, Arena R, Sietsema K, Myers J, Coke L, Fletcher GF, et al. Clinician’s Guide to Cardiopulmonary Exercise Testing in Adults. Circulation. 2010;122(2):191-225.

2.	Stanojevic S, Kaminsky DA, Miller MR, Thompson B, Aliverti A, Barjaktarevic I, et al. ERS/ATS technical standard on interpretive strategies for routine lung function tests. Eur Respir J. 2022;60(1).

3.	Thomsen LP, Karbing DS, Smith BW, Murley D, Weinreich UM, Kjærgaard S, et al. Clinical refinement of the automatic lung parameter estimator (ALPE). J Clin Monit Comput. 2013;27(3):341-50.

4.	Rees SE, Kjaergaard S, Perthorgaard P, Malczynski J, Toft E, Andreassen S. The automatic lung parameter estimator (ALPE) system: non-invasive estimation of pulmonary gas exchange parameters in 10-15 minutes. J Clin Monit Comput. 2002;17(1):43-52.

5.	Lang RM, Badano LP, Mor-Avi V, Afilalo J, Armstrong A, Ernande L, et al. Recommendations for Cardiac Chamber Quantification by Echocardiography in Adults: An Update from the American Society of Echocardiography and the European Association of Cardiovascular Imaging. Journal of the American Society of Echocardiography. 2015;28(1):1-39.e14.

6.	Rako ZA, Yogeswaran A, Lakatos BK, Fábián A, Yildiz S, da Rocha BB, et al. Clinical and functional relevance of right ventricular contraction patterns in pulmonary hypertension. J Heart Lung Transplant. 2023;42(11):1518-28.

7.	Rako ZA, Kremer N, Yogeswaran A, Richter MJ, Tello K. Adaptive versus maladaptive right ventricular remodelling. ESC Heart Fail. 2023;10(2):762-75.

8.	Bauer P, Kraushaar L, Most A, Hölscher S, Tajmiri-Gondai S, Dörr O, et al. Impact of Vascular Function on Maximum Power Output in Elite Handball Athletes. Res Q Exerc Sport. 2019;90(4):600-8.
